# Immunogenicity and safety of fractional doses of yellow fever vaccines: a randomized, double-blind, non-inferiority trial

**DOI:** 10.1101/2020.08.18.20177527

**Authors:** Aitana Juan-Giner, Derick Kimathi, Kyra H. Grantz, Mainga M. Hamaluba, Patrick Kazooba, Patricia Njuguna, Gamou Fall, Moussa Dia, Ndeye S. Bob, Thomas P. Monath, Alan D. Barrett, Joachim Hombach, Edgar M. Mulogo, Immaculate Ampeire, Henry K. Karanja, Dan Nyehangane, Juliet Mwanga-Amumpaire, Derek A.T. Cummings, Philip Bejon, George M. Warimwe, Rebecca F. Grais

**Author notes:** Corresponding author: Rebecca F Grais; Address: 34 Avenue Jean Jaurès, 75019 Paris, France. Co-first author (Aitana Juan-Giner, Derick Kimathi). Co-senior author (George M. Warimwe, Rebecca F. Grais).

## Abstract

**Background:** Yellow fever vaccine stocks have been insufficient to cover exceptional demands for outbreak response. Fractional dosing evidence is limited to the 17DD substrain vaccine. We assessed the immunogenicity and safety of one-fifth fractional dose compared to standard dose of each of the four WHO-prequalified yellow fever vaccines produced from three substrains.

**Methods:** We conducted a randomized, double-blind, non-inferiority trial in Mbarara, Uganda and Kilifi, Kenya. 960 participants aged 18-59 years without history of yellow fever vaccination or infection were recruited from communities and randomized to a vaccine and dosage. Vaccine was administered subcutaneously by unblinded nurse. Other study personnel and participants were blinded to vaccine allocation. Primary immunogenicity outcome, seroconversion, was measured in the per-protocol population; safety outcomes included all vaccinated participants. We defined non-inferiority as less than 10% decrease in seroconversion in fractional compared to standard dose arms 28 days post-vaccination. Seroconversion was ≥4-fold rise in neutralizing antibody titers measured by 50% plaque reduction neutralization test.

ClinicalTrials.gov Identifier: NCT02991495, completed.

**Findings:** Between 6^th^ November 2017 and 21^st^ February 2018, 960 participants, total sample goal, were randomized. The primary per-protocol analysis includes 899 participants, 110 to 117 participants per arm. The absolute difference in seroconversion between fractional and standard doses by vaccine was 1.71% (95%CI: −2.60, 5.28) for Bio-Manguinhos, −0.90% (95%CI: −4.24, 3.13) for Chumakov Institute, 1.82% (95%CI: −2.75, 5.39) for Institut Pasteur de Dakar, 0.0% (95%CI: −3.32, 3.29) for Sanofi Pasteur. Fractional doses from all four vaccines met the non-inferiority criterion. The most common related AEs were headache (22.2%), fatigue (13.7%), myalgia (13.3%) and self-reported fever (9.0%). There were no safety concerns.

**Interpretation:** These results support fractional dosing of all WHO-prequalified yellow fever vaccines for the general adult population for outbreak response in situations of vaccine shortage.

**Funding:** The study was funded by Médecins Sans Frontières Foundation, Wellcome Trust (grant no. 092654) and the UK Department for International Development. Vaccines were donated in kind.

## Introduction

Yellow fever is a mosquito-borne viral disease endemic in 44 countries with periodic outbreaks (1). Four live attenuated yellow fever virus vaccines derived from multiple strains of 17D are World Health Organization (WHO)-prequalified, including 17DD vaccine from Bio-Manguinhos/Fiocruz (Brazil), 17D-213 vaccine from Federal State Unitary Enterprise of Chumakov Institut of Poliomyelitis and Viral Encephalitides (IPVE) (Russia), and 17D-204 vaccine from Institut Pasteur de Dakar (Senegal) and Sanofi Pasteur (France). All have been widely used and are considered safe and effective (1). WHO recommends routine vaccination in all countries where the disease is endemic, vaccination of travelers to those areas, and mass vaccination for prevention or control of outbreaks. A stockpile of 2 million doses was reserved for outbreak response in 2000 and increased to 6 million in 2014 to facilitate response to major multi-country, but unpredictable outbreaks (2).

An outbreak of yellow fever in 2016 in Angola raised major concerns about the adequacy of vaccine supply. Routine vaccination was suspended in some African countries to meet demand during the Angola outbreak (3) and a subsequent outbreak in Democratic Republic of Congo (DRC) further contributed to a global shortage (4). In response, WHO reviewed the evidence on fractional dosing of yellow fever vaccine as a dose-sparing option and exceptionally recommended considering off-label use of fractional doses, administered by the standard subcutaneous or intramuscular route, to extend supply in response to ongoing outbreaks (4,5). Fractional doses (defined as one-fifth of the standard dose) of the yellow fever vaccine produced by Bio-Manguinhos/Fiocruz (17DD substrain) were given to approximately 7.5 million non-pregnant adults and children ≥2 years of age in Kinshasa, DRC in August, 2016 (6). Fractional dosing was again used in response to outbreaks in 2017-2018 in the states of São Paulo, Rio de Janeiro and Bahia, Brazil to vaccinate almost 17 million people (7). Four studies have assessed the immunogenicity of fractional doses of yellow fever vaccine. One of these studies (8) used an old vaccine formulation no longer produced and another focused on the intradermal administration (9). The evidence base to support WHO recommendations was primarily from a dose-finding study of the 17DD substrain vaccine amongst healthy male army recruits in Brazil that showed seroconversion rates greater than 97% with doses as low as 587 IU/dose (10) and similar virological and immunological kinetics with doses down to 3,013 IU/dose compared to the released dose of 27,476 IU/dose (11). Based on historic data, standard doses of yellow fever vaccines should contain not less than 1,000 IU/dose (12). However, release specifications vary by manufacturer, related to the stability of the vaccine, and are generally several-fold higher than the minimum specification (13). WHO recommended that the minimal dose administered, standard or fractional, should preferentially contain 3,000 IU/dose, hence the decision on the dose fractioning (e.g. one-half or one-fifth of the standard dose) should consider the potency of the vaccine batch (4). The dose-finding study conducted in Brazil (10,11) provides information for only one of the WHO-prequalified vaccines produced, that of the 17DD substrain. During the fractional dose campaign in Kinshasa, DRC, a study confirmed the immunogenicity of fractional doses of the 17DD substrain vaccine in a large-scale campaign (14). Information is lacking for the remaining substrains.

We assessed the immunogenicity and safety of fractional doses of all four WHO-prequalified yellow fever vaccines. This is the first clinical trial where all four WHO-prequalified vaccines have been used in the same study, providing information on the different substrains. This study fills a critical gap to provide evidence to broaden recommendations to consider including use of all WHO-prequalified yellow fever vaccines for fractional dosing (4).

## Methods

### Study design

We conducted a two-center, double-blind, individually-randomized trial in Mbarara, Uganda and Kilifi, Kenya (15). The study took place at the Epicentre Mbarara Research Centre in Mbarara, Uganda and the Kenya Medical Research Institute (KEMRI)-Wellcome Trust Research Programme clinical trials facility in Kilifi, Kenya. Mbarara district is located in proximity to Masaka, Rukungiri, and Kalangala districts which registered confirmed yellow fever cases in 2016 (16).

The study protocol was approved by the ethics committee of the World Health Organization (Switzerland); Scientific & Ethics Review Unit, Kenya Medical Research Institute (Kenya); Oxford Tropical Research Ethics Committee (United Kingdom); Mbarara University of Science and Technology Research Ethics Committee (Uganda); and the Uganda National Council of Sciences and Technology (Uganda). Approval was obtained from the national regulatory authorities in Uganda and Kenya. The trial was conducted in accordance with Good Clinical Practice guidelines.

### Participants

Participants were recruited from rural communities in the Mbarara municipality, Mbarara district, Uganda and Kilifi County, Kenya. Communities were informed about the trial using locally adapted strategies. Individuals interested in participating were invited to the study sites. Written informed consent was required to participate.

Individuals were eligible to participate if they were 18 to 59 years of age; had no contraindications for vaccination; were not pregnant or lactating; had no history of previous yellow fever vaccination or infection; did not require yellow fever vaccination for travel; and were able to comply with study procedures.

### Randomization and masking

Participants were individually randomized to one of 8 arms corresponding to the four prequalified yellow fever vaccines at standard or fractional dose. Unique allocation numbers were prepared by an independent statistician (DiagnoSearch LifeSciences, Mumbai, India) using a computer-generated random number list with non-disclosed fixed blocks of size 10 with equal allocation to dosage within a block and to a given manufacturer by site. The allocation sequence was concealed using pre-prepared, sequentially numbered scratch-off booklets stored under key. After enrollment, the vaccination nurse scratched off the randomization code indicating the allocated vaccine and dosage for the participant. Vaccines were reconstituted and administered in a private room not accessible to other study staff.

Participants were blinded by masking the volume of the syringe with opaque tape. The vaccination nurse and supervisor overseeing vaccination were aware of allocation; personnel assessing outcomes and investigators were blinded to vaccine and dosage throughout.

### Procedures

A batch of yellow fever vaccine of standard 10-dose vials was selected from each manufacturer with potency at time of release closest to internal minimum specification. Vaccine potencies were independently measured at the National Institute for Biological Standards and Control (United Kingdom) (Table 1). The freeze-dried preparations and diluents were kept at 2-8°C until administration. Each day, after a participant was randomized to a specific vaccine, a vaccine vial was reconstituted using the manufacturer’s diluent. A syringe to administer fractional or standard doses was prepared immediately prior to vaccination. Reconstituted vaccines were kept in a vaccine carrier at 2-8°C per WHO and manufacturer requirements. Any remaining reconstituted vaccine was discarded after 6 hours. Fractional doses consisted of one-fifth (0.1ml) of the standard dose (0.5ml). Vaccine was administered subcutaneously in the deltoid region using 0.5ml auto-disable syringes (needle size 25G x 3/4”) with a 45° injection angle for the standard 0.5ml dose and 0.1ml auto-disable syringes (needle size 26G x 3/8”) at 90° injection angle for the 0.1ml fractional dose.

**Table 1.** Characteristics of study vaccines

| **Manufacturer** | **Substrain** | **Expiry date** | **IU/dose Mean (SD)** |
| --- | --- | --- | --- |
| Bio-Manguinhos/Fiocruz (Bio-Manguinhos) | 17DD | 31/10/2018 | 38,905 (1.26) |
| Federal Stata Unitary Enterprise of Chumakov Institute of Poliomyelitis and Viral Encephalitides (Chumakov IPVE) | 17D-213 | 28/02/2018 | 43,652 (1.12) |
| Institut Pasteur de Dakar (IPD) | 17D-204 | 31/03/2019 | 6,761 (1.74) |
| Sanofi Pasteur | 17D-204 | 28/02/2019 | 16,596 (1.26) |

Note: Standard doses of the vaccines were tested at the National Institute for Biological Standards and Control (NIBSC), UK, December 2017. The IU/dose is the mean of 3 assays; SD: Standard deviation.

Participants were followed at 10 (±1) days, 28 (±3) days, and 365 (±14) days post-vaccination. At each study visit, participants had a medical consultation and a blood sample was obtained at the initial visit prior to vaccination and subsequently at each scheduled follow-up visit.

Serum samples were separated and aliquoted within 4 hours of collection and stored in −80°C freezers at each site. Serum samples were analyzed at the Institut Pasteur de Dakar (Senegal), where neutralizing antibody titers against yellow fever were assessed by 50% plaque reduction neutralization test (PRNT_50_) as primary outcome and PRNT_90_ as supplementary (17,18). Laboratory personnel were blinded to vaccine and dose allocation.

The attenuated yellow fever vaccine strain 17D-204 sourced by Institut Pasteur de Dakar was used as challenge strain for the neutralization assays. Viral stock preparation was in C6/36 cells and titrated by plaque assay method previously described (19). Briefly, a defined virus concentration of 10^3^ plaque forming units per ml and diluted and heat-inactivated sera were titrated in serial two-fold dilutions from 1:10 to 1:20480. Serum and virus were mixed and incubated before overlaying with 0.6% carboxy-methyl-cellulose and 3% fetal calf serum in L-15 Leibovitz’s media. Following 4 to 5 days of incubation at 37°C, plaques were counted, and antibody titer was determined as the dilution that reduced observed plaques by 50% and 90% compared to the number observed in control wells.

### Outcomes

The primary outcome is non-inferiority in the proportion of participants who seroconverted, as determined by neutralization assay (PRNT_50_), by day 28 post-vaccination for fractional dose compared to standard dose for each WHO-prequalified vaccine. Seroconversion is defined as a ≥4-fold rise in neutralizing antibody titer between pre-vaccination and post-vaccination samples.

Secondary outcomes at day 28 post-vaccination include assessment of geometric mean titers (GMT) and geometric mean fold increase (GMFI) (i.e. geometric mean of the ratios of post-vaccination titer to pre-vaccination titer). Seroconversion, GMT, and GMFI were also assessed at 10 days and 1 year after vaccination to assess the rapidity of protection as well as the lasting effect of vaccination.

Safety outcomes include assessment of adverse events (AEs) for 28 days after administration of fractional and standard doses and serious adverse events (SAEs) throughout the duration of follow-up. SAEs were defined as any new health-related problem that resulted in death, was life-threatening, necessitated hospitalization or prolongation of existing hospitalization, or resulted in disability or incapacity, that occurred during the study follow-up. AEs included all untoward medical events and were evaluated in the 30 minutes following vaccination and up to 28 days post-vaccination. At the 10- and 28-days post-vaccination visits, a clinician asked for the presence of local reaction, headache, fatigue, muscle pain, fever, gastrointestinal problem and any other symptom since the previous visit. Participants were also asked to report any other symptoms or concerns during and outside scheduled visits. The nature, relatedness, severity and outcome of each AE were recorded (20,21). All events were coded using MedDRA dictionary, version 20.0.

### Statistical analysis

Weighing the public health consequence of loss of protection against an increase in available vaccine doses and coverage, non-inferiority of the fractional dose was defined as seroconversion rate at 28 days post-vaccination no more than 10% lower than the standard dose. We assumed 95% seroconversion with standard doses and considered that a loss of protection to 85% seroconversion with fractional doses would still ensure protection above 80% necessary to interrupt local transmission (22). The non-inferiority margin is supported by a modelling study where fractional doses were shown beneficial in high-transmission areas if 80 to 90% efficacious (23). To detect a non-inferiority margin of −10% in seroconversion 28 days post-vaccination, with a 2.5% level of significance for a one-sided test, 90% power, and accounting for 5% loss to follow-up and 15% baseline yellow fever seropositivity (i.e.PRNT_50_ titer ≥10), a sample size of 120 per manufacturer and dose was required, for a total of 240 per manufacturer. In total, 960 participants were to be recruited (480 per study site).

The primary analysis was non-inferiority in the proportion of participants seronegative to yellow fever at baseline (i.e. PRNT_50_ titer <10) who seroconvert at 28 days post-vaccination for the fractional dose compared to the standard dose of each WHO-prequalified vaccine. We defined seroconversion as ≥4-fold rise in PRNT_50_ titer between baseline pre-vaccination and post-vaccination visits. Secondary endpoints include assessment of geometric mean PRNT_50_ titers (GMT) and geometric mean fold increase (GMFI) at 28 days post-vaccination, and seroconversion, GMT and GMFI at 10 days and one year after vaccination. Analysis using PRNT_90_ titers are presented in the supplementary material.

Analysis consisted of pairwise comparisons of fractional versus standard dose of the same manufacturer’s vaccine. No comparisons were made of vaccines from different manufacturers. Comparison of baseline characteristics were done using χ^2^ tests (or Fisher’s exact test for smallest count <5) for categorical variables and Student’s t-test for continuous variables. Any PRNT_50_ titer reported as seronegative (below the Limit of Quantification (LOQ), <10) was converted to LOQ/2. Any PRNT_50_ titer >20480 was assigned as 20480.

The number and percentage of participants who seroconverted are presented by study arm (i.e. manufacturer and dose) with two-sided exact 95% confidence interval (CI) based on the Clopper-Pearson method. Non-inferiority for the primary outcome was assessed by constructing a two-sided 95% CI using the Wilson score interval of the point difference between seroconversion rates in the fractional and standard dose arms. Fractional doses were considered non-inferior if the lower bound of the CI for difference in seroconversion was greater than −10%.

Two-sided 95% CIs of the mean difference between log GMT and log GMFI between the standard and fractional dose of each vaccine were generated using the t-distribution. Intervals were transformed to show the ratio of GMT and GMFI for the fractional compared to standard dose.

Immunogenicity outcomes were assessed in the per-protocol population (PP) and the intent-to-treat population (ITT). The PP population included all participants for whom the eligibility criteria were appropriately applied, who were seronegative at baseline (PRNT_50_ below LOQ), and who had a PRNT_50_ titer at the time point of interest. The ITT population included any vaccinated participant with at least one PRNT_50_ result post-vaccination.

AEs and SAEs were summarized as number and percentage by study arm. Safety outcomes were assessed in all vaccinated participants.

Data analysis was conducted in R, version 3.6.1. A data safety monitoring board (DSMB) regularly reviewed study data.

ClinicalTrials.gov Identifier: NCT02991495

## Results

From 6^th^ November 2017 to 21^st^ February 2018, 1,029 participants were assessed for eligibility, 69 were ineligible, and 960 were randomized to a vaccine manufacturer and dosage (120 per study arm) and vaccinated (Figure 1). Nearly all (99.2%) participants completed the day 28 post-vaccination visit and 96.8% completed the one year post-vaccination visit. The most frequent reasons for discontinuation were migration out of study area (n=13), loss to follow-up (n=10) and non-compliance with study visits (n=4). There were 2 discontinuations due to protocol violation: inclusion of a 65-year-old man and re-vaccination of a participant attending the day 28 post-vaccination visit.

**Figure 1.**
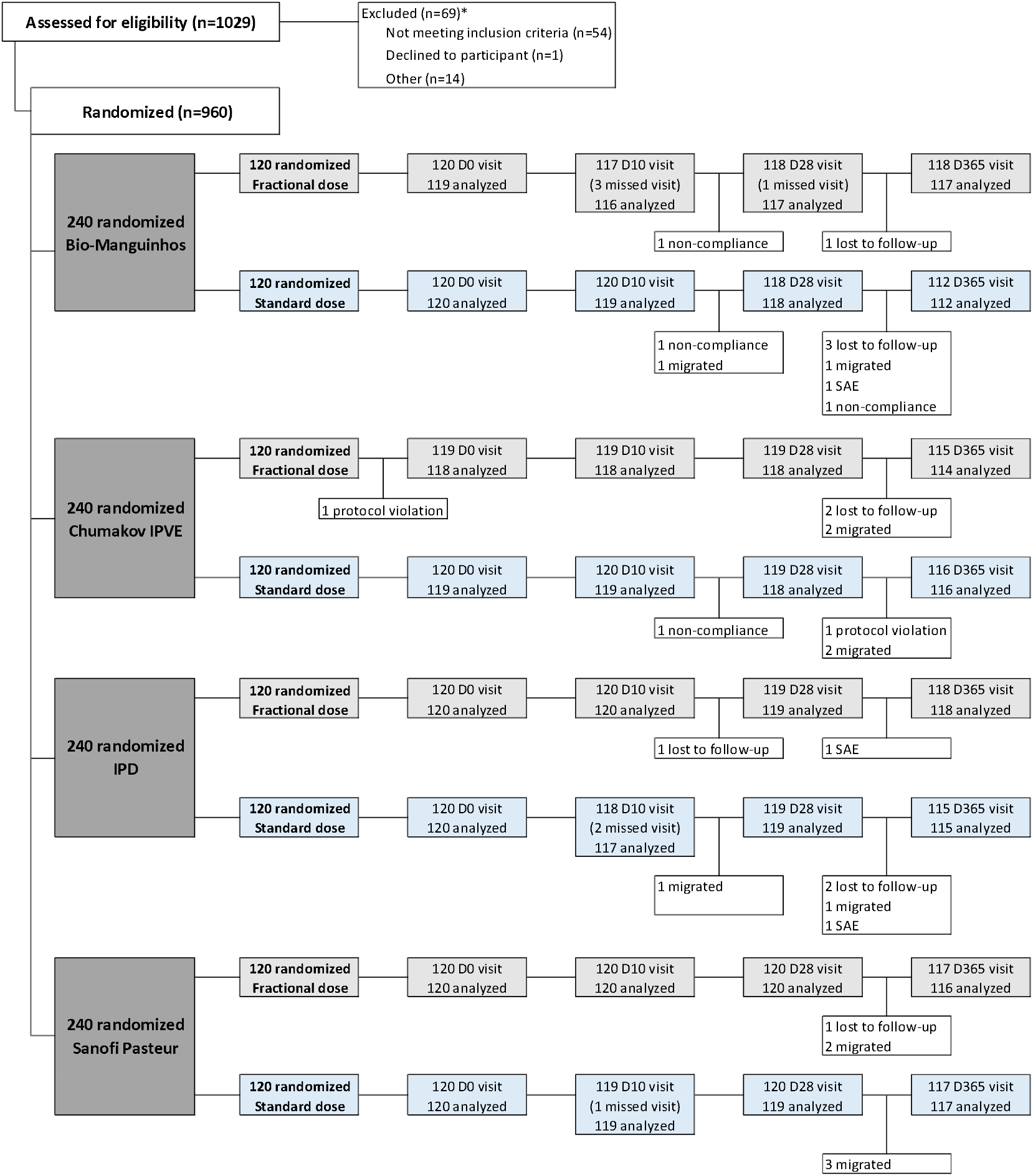
Trial profile. * Participants deemed ineligible include: Immunodeficiency (n=21), new HIV diagnosis or in need of referral to HIV treatment (n=13) pregnant/lactating (n=9), allergy to egg proteins (n=5), history of yellow fever vaccination (n=3), unable to complete follow-up (n=2), acute febrile illness (n=1), requiring vaccination for travelling (n=1). Other criteria: underlying diseases (n=3) and incomplete screening (n=11). Participants could fail to meet more than one eligibility criterion. Note: number analyzed refers to the number included in the immunogenicity analysis at each time point. Differences with number of completed visits are due to missing PRNT result.

The primary analysis (PP) included 110 to 117 participants per arm, 93.6% of the randomized participants. There were 49 participants with detectable PRNT_50_ at baseline (included in ITT analysis) and 11 participants with missing PRNT_50_ results at baseline or 28 days follow-up (Figure 1).

The mean age of participants was 35.7 years of age and 55.1% were females. Participant characteristics were similar at baseline between study arms (Table 2).

**Table 2.** Baseline demographic and clinical characteristics

|  | **Bio-Manguinhos** |  | **Chumakov IPVE** |  | **IPD** |  | **Sanofi Pasteur** |  |
| --- | --- | --- | --- | --- | --- | --- | --- | --- |
|  | Fractional<br>(N=120) | Standard<br>(N=120) | Fractional<br>(N=119) | Standard<br>(N=120) | Fractional<br>(N=120) | Standard<br>(N=120) | Fractional<br>(N=120) | Standard<br>(N=120) |
| Age at enrolment, Mean (SD) | 34.5<br>(11.5) | 35.0<br>(11.1) | 37.2<br>(11.4) | 35.9<br>(11.8) | 35.5<br>(11.4) | 35.4<br>(11.7) | 35.3<br>(11.7) | 36.8<br>(12.9) |
| Female sex, no (%) | 73 (60.8) | 73<br>(60.8) | 58 (48.7) | 62<br>(51.7) | 64 (53.3) | 72<br>(60.0) | 61(50.8) | 66<br>(55.0) |
| HIV+, no (%) | 4(3.3) | 1 (0.8) | 0 (0) | 0 (0) | 1 (0.8) | 1 (0.8) | 1 (0.8) | 2 (1.7) |
| Seropositive to YF at baseline, no (%) | 6* (5.0) | 1 (0.8) | 7** (5.9) | 4* (3.4) | 7 (5.8) | 9 (7.5) | 7 (5.8) | 8 (6.7) |

\*N=119; \*\*N=118 due to missing PRNT titer at baseline
Seropositive to YF at baseline defined as PRNT<sub>50</sub> titer ≥10

At 28 days post-vaccination, seroconversion rates were high in all arms, with at least 98.2% of participants seroconverting (Table 3). The difference in seroconversion between fractional and standard dose arms was 1.71% (95%CI: −2.60, 5.28) for the vaccine produced by Bio-Manguinhos/Fiocruz with the 17DD substrain, −0.90% (95%CI: −4.24, 3.13) for the vaccine produced by Chumakov IPVE with the 17D-213 substrain, 1.82% (95%CI: −2.75, 5.39) for the vaccine produced by Institut Pasteur Dakar and 0.0% (95%CI: −3.32, 3.29) for the vaccine produced by Sanofi Pasteur, both derived from the 17D-204 substrain. The lower bound of the 95% confidence interval for the difference in seroconversion between fractional and standard dose arms for each vaccine manufacturer excluded the defined non-inferiority margin of - 10%, indicating non-inferiority of the fractional dose for each vaccine (Table 3 and Figure 2). Non-inferiority is also met considering PRNT_90_ results (Table S1). Results for the ITT population at 28 days post-vaccination were similar, with a lower bound of the confidence interval for the difference in seroconversion between fractional and standard dose arms for each vaccine manufacturer of −2.58%, −6.11%, −2.11% and −3.10% (Table S2). Seroconversion was high even among those adults with neutralizing antibodies against yellow fever at baseline (Table S3).

**Table 3.**
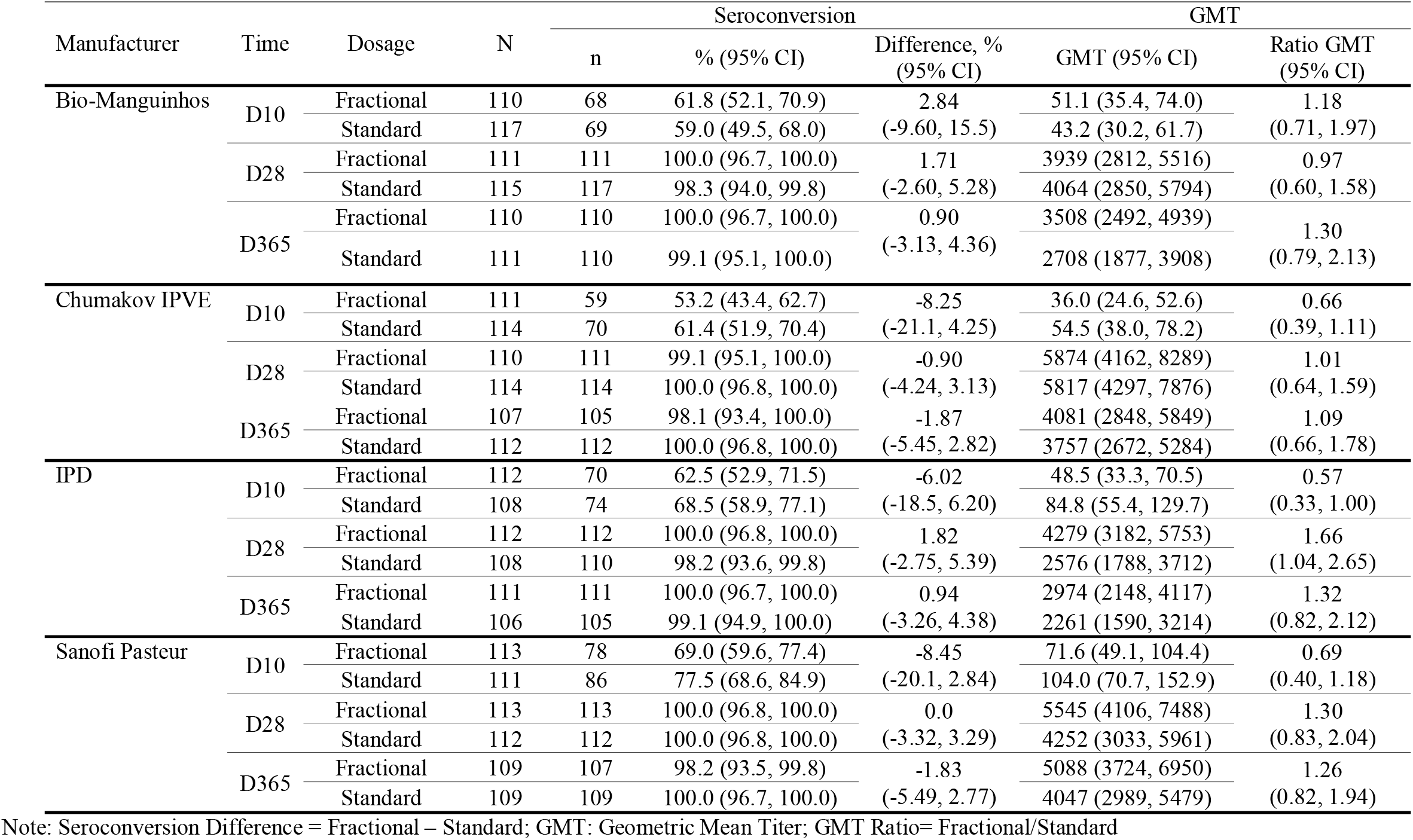
Non-inferiority of seroconversion and GMT by PRNT_50_ in fractional vs. standard dose of yellow fever vaccine at Day 10, Day 28 and y 365 post-vaccination, PP population, by vaccine manufacturer

At 10 days post-vaccination, the percentage of participants that seroconverted in each arm varied from 53.2% to 77.5%, with no differences between fractional and standard doses by vaccine manufacturer considering the large and overlapping confidence intervals (Table 3). Seroconversion rates remained high one year after vaccination, with between 98.1% and 100% of those who received a fractional dose and >99% of those who received the standard dose maintaining a ≥4-fold rise in PRNT_50_ titer from baseline (Table 3).

GMTs of neutralizing antibodies 28 days post-vaccination were high in each arm, with titers ranging from 3939 to 5874 in the fractional dose arms and 2576 to 5817 in the standard dose arms. For one vaccine GMTs were higher (Table 3). At 10 days post-vaccination, GMTs were lower compared to 28 days post-vaccination and were, on average, lower in the fractional dose compared to the standard dose arm for three vaccines, though the confidence intervals for the difference are wide and overlap 0 for all but one vaccine (Table 3). At one year post-vaccination, GMTs remained high in all study arms and were consistently higher in the fractional dose arms (range: 2974-5088) compared to standard dose arms (range: 2261-4047). There was some evidence for a slow decline in GMT from 28 days to one year post-vaccination in all arms (Figure S5).

GMFIs showed an increase in neutralizing antibody titers from baseline at each time point for all vaccine arms, with GMFIs reaching highest point 28 days post-vaccination (Tables S6, S10 and S11).

AEs were common with between 58% to 63% of participants per arm reporting at least one AE and 32 to 49% reporting an AE classified as vaccine-related within 28 days of vaccination. There were similar number of events across all arms. The most common related AEs were headache (22.2%), fatigue (13.7%), myalgia (13.3%), and self-reported fever (9.0%) (Table 4 and Table S13).

**Table 4.** Occurrence of most common Adverse Events up Day 28 post-vaccination by MedDRA (v20.0) Preferred Term, all vaccinated participants.

|  | Bio-Manguinhos |  | Chumakov IPVE |  | IPD |  | Sanofi Pasteur |  |
| --- | --- | --- | --- | --- | --- | --- | --- | --- |
| *No. $\geq 1$ event (%)* | Frac.<br>N=120 | Stand.<br>N=120 | Frac.<br>N=119 | Stand.<br>N=120 | Frac.<br>N=120 | Stand.<br>N=120 | Frac.<br>N=120 | Stand.<br>N=120 |
| Headache | 21<br>(17.5) | 26<br>(21.7) | 27<br>(22.7) | 30<br>(25.0) | 30<br>(25.0) | 22<br>(18.3) | 29<br>(24.2) | 28<br>(23.3) |
| Fatigue | 8 (6.7) | 15<br>(12.5) | 18<br>(15.1) | 12<br>(10.0) | 19<br>(15.8) | 21<br>(17.5) | 21<br>(17.5) | 17<br>(14.2) |
| Myalgia | 15<br>(12.5) | 18<br>(15.0) | 14<br>(11.8) | 15<br>(12.5) | 14<br>(11.7) | 22<br>(18.3) | 14<br>(11.7) | 16<br>(13.3) |
| Pyrexia | 9 (7.5) | 11 (9.2) | 12<br>(10.1) | 9 (7.5) | 12<br>(10.0) | 9 (7.5) | 10 (8.3) | 14<br>(11.7) |
| Abdominal pain | 4 (3.3) | 3 (2.5) | 3 (2.5) | 8 (6.7) | 9 (7.5) | 1 (0.8) | 6 (5.0) | 3 (2.5) |

A table with all reported AEs up to Day 28 post-vaccination by MedDRA coding, study arm and vaccine manufacturer, is presented in the supplementary material.

In total, 9 SAEs, including 3 deaths, were reported. All were classified as not related to the study vaccines (Tables S18 and S19).

## Discussion

This study shows that one-fifth fractional doses of the four WHO-prequalified yellow fever vaccines met the main outcome of non-inferiority in seroconversion 28 days post-vaccination by PRNT_50_ compared to the standard dose. While not firmly established, non-human studies suggest that neutralizing titers ≥40-50 protect against lethal yellow fever infection, and these levels have been used as the cut-off for protection from infection in humans (24). At 28 days post-vaccination, almost all participants showed high levels of neutralizing antibodies far in excess of the assumed protective threshold. Seroconversion rates and GMT remain high up to one year post-vaccination for both fractional and standard doses for all vaccines. There were no major safety concerns with either the standard or fractional dose of each of the four WHO-prequalified vaccines.

The results of our trial are aligned with previous studies on fractional dosing using the 17DD substrain vaccine in other contexts showing that nearly all vaccinated individuals seroconvert within 28 days of vaccination (10,14). We now extend the evidence base to randomized comparisons for all WHO-prequalified vaccines and to a general adult population in rural sub-Saharan Africa. Previous studies indicate that the immune response generated by fractional doses is long-lived (14,25,26), consistent with our findings of a robust immunogenic response at one year post-vaccination, though longer-term studies are warranted.

We assessed, for the first time in a randomized trial including all four WHO-prequalified yellow fever vaccines, seroconversion rates at 10 days post-vaccination in all participants. These results show overall lower seroconversion and GMTs in the fractional dose arms, suggesting a possible delayed immune response of the fractional doses. However, previous studies have shown that 80-90% of standard dose vaccine recipients have protective levels of neutralizing antibodies 10 days after vaccination (27). This proportion is lower in our study, even among participants that received a standard dose of vaccine. Our study was performed in an African population, which may have lower B and T cell responses to yellow fever vaccination than European populations (28). Given the likely use of fractional doses in outbreak responses, the low rates of seroconversion at 10 days are concerning and emphasize the need for early vaccination campaigns in outbreak response. Current International Health Regulations, which allow for travel 10 days post-vaccination, may require further study to fully understand implications for standard dose vaccination and any future recommendations for fractional doses.

The results of this study are subject to several limitations. First, it is important to highlight that the study was insufficiently powered to assess non-inferiority of the fractional doses at 10 days post-vaccination. Overall, 67% of participants seroconverted in the standard dose arms at 10 days post-vaccination. Due to the low rates of seroconversion our study has only 35% power to detect a −10% non-inferiority margin difference between fractional and standard dose groups at 10 days post-vaccination. This is insufficient to draw conclusions on non-inferiority and additional studies could be warranted to better understand the early immune response to vaccination with fractional doses. Second, we did not assess for presence of neutralizing antibodies against other flaviviruses that could potentially interfere with the response to yellow fever vaccine. To compensate for this, participants were asked about history of infection with Zika, Dengue or West Nile virus, but only one participant reported being aware of a previous infection. In total, 49 participants had neutralizing antibodies against yellow fever at baseline. This represents a very low number. These participants were excluded from the PP analysis. We conducted supplementary analyses at day 10, day 28 and day 365 including participants who were seropositive at baseline and found no difference in interpretation.

Finally, the primary limitation for the generalizability of results are the vaccines used in the study. In this study, we used vaccines as close as possible to each manufacturer’s internal minimum specification for potency. Vaccines titers were, however, high, with potencies between 6 and 43 times the minimum specification established by WHO. All fractional doses still contained potencies above the minimum specification of 1000 IU. Vaccines also differed in expiry dates and had between 3 and 16 months of remaining shelf-life (i.e. between 1/8^th^ and less than 1/5^th^ remaining shelf-life) at the time of testing at NIBSC (Table 1). Current yellow fever vaccines were developed more than 80 years ago (1) following a production process that has not significantly changed since and lacking clinical data to support minimum specifications (13). Historically, vaccines have been released at higher titers than the recommended minimum, partly to account for the loss of potency during shelf-life (13). Average doses of currently produced WHO-prequalified vaccines vary between 12,874 and 43,651 IU and consequently, titers of fractional doses would in principle still exceed WHO’s minimum specification (29). We consider that the batches selected for this study represent the WHO-prequalified vaccines as currently produced by the different manufacturers and that results should be generalizable to other batches as long as there is not a significant change in the manufacturing process. Therefore, our study provides additional confidence in using fractional doses for the vaccines as currently produced.

Given the longstanding practice of releasing batches with high potency, and the dependence on historical precedent for dose requirements, there was significant uncertainty regarding the use of fractional dosing, even for doses above the minimum specifications. This is reflected in the WHO recommendations for fractional dosing that the minimal dose administered should preferentially contain 3000 IU/dose, three times the minimum specification for vaccine potency (4). Our data suggest that, even at the lower end of the range of vaccine potency and with minimal remaining shelf-life, fractional doses can be an option for outbreak response.

These results support the use of one-fifth fractional doses of the four WHO-prequalified yellow fever vaccines for the general adult population when there are insufficient standard doses to protect the population at risk during an outbreak, thereby filling a critical knowledge gap to support WHO policy on the use of fractional dosing of yellow fever vaccine (5). The use of fractional dosing can expand the outbreak stockpile up to five-fold and therefore will be a critical back-up tool in case of vaccine supply shortage during yellow fever outbreak response. We are currently conducting sub-studies to assess immunological non-inferiority and safety of fractional doses among children from 9 months to 5 years of age and adults living with HIV.

## Data Availability

Analytic code and a deidentified dataset are available upon request to.

## Contributors

AJG, PB, KHG, DC, JH, TM, AB and RFG designed the study; PK, DK and GW collected the data; KHG conducted the statistical analysis, GF, NSM and MD conducted the laboratory analyses; and AJG prepared the first draft of the manuscript. AJG and RFG vouch for the accuracy and completeness of the data and analyses reported. All authors contributed to the interpretation of data, critically reviewed the manuscript and decided to publish the paper.

## Disclaimer

JH is staff of the World Health Organization. The findings and conclusions in this report are those of the authors and do not necessarily represent the official position of the World Health Organization

## Data sharing

Data collected for the study, including deidentified participant data, data dictionary and additional related documents such as study protocol and statistical analysis plan, will be made available to others upon request to, following Epicentre’s data sharing policy and in accordance with WHO statement on public disclosure of clinical trial results.

## Acknowledgements

The study was funded by Médecins Sans Frontières Foundation, Wellcome Trust (grant no. 092654) and the UK Department for International Development. The vaccines were donated in kind by Bio-Manguinhos/Fiocruz (Brazil), Federal State Unitary Enterprise of Chumakov Institut of Poliomyelitis and Viral Encephalitides (Russia), Institut Pasteur de Dakar (Senegal), and Sanofi Pasteur (France). Epicentre receives core funding from Médecins Sans Frontières.

We wish to thank study participants and the Scientific Committee, the DSMB and the National Institute for Biological Standards and Control (United Kingdom) for critical support to this project. We also thank the YEFE teams in KEMRI-Wellcome Trust Research Programme, Kilifi, Kenya and in Epicentre Mbarara Research Centre, Mbarara, Uganda.

## Research in context

### Evidence before this study

In July 2016, following major yellow fever (YF) outbreaks in two countries, WHO published a secretariat information paper including a review of studies assessing the immunogenicity of fractional doses of YF vaccines and recommended consideration of fractional doses to manage a vaccine shortage. Following this, fractional doses of YF vaccine produced by Bio-Manguinhos/Fiocruz (17DD substrain) were given to approximately 7.5 million non-pregnant adults and children ≥2 years of age in Kinshasa, Democratic Republic of Congo. The evidence base to support this action was limited to a single vaccine substrain and to a specific context. To broaden and simplify recommendations, WHO called for additional research to be conducted. We designed a trial to assess non-inferiority in seroconversion of fractional (one-fifth dose) versus standard dose for each of the four WHO-prequalified YF vaccines at 28 days post-vaccination in an adult population in Kenya and Uganda. We selected vaccine batches as close as possible to each manufacturer’s minimum release specification.

### Added value of this study

This is the first randomized controlled trial assessing all four WHO-prequalified YF vaccines, providing information on the immunogenicity and safety of fractional doses of the different vaccine substrains at 10 days, 28 days and one year post-vaccination. The results show that, at 28 days post-vaccination, most participants had high levels of neutralizing antibodies and that seroconversion rates in the fractional dose arms were non-inferior to standard dose for each of the four vaccines. Seroconversion rates and neutralizing antibodies remained high up to one year post-vaccination for both fractional and standard doses for all vaccines. These results are aligned with previous studies using the 17DD substrain vaccine but extend the evidence to randomized comparisons of all four vaccines and to a sub-Saharan Africa context.

### Implications of all the available evidence

Our study supports the use of one-fifth fractional doses of all four WHO-prequalified yellow fever vaccines for the general adult population and fills a critical knowledge gap to support WHO policy on the use of fractional dosing of yellow fever vaccine for outbreak response. The immunogenicity and safety of fractional dosing in children and specific populations, such as those living with HIV, is yet to be determined. Long-term studies are warranted to confirm the duration of protection.

