## Supplementary Appendix for "Immunogenicity and safety of fractional doses of yellow fever vaccines: a randomized, double-blind, non-inferiority trial"

**Immunogenicity Analysis**

Immunogenicity assessment

Viral stock preparation was done in C6/36 cells and titrated by plaque assay method. The attenuated yellow fever vaccine strain 17D is used as challenge strain for the neutralization assays. The yellow fever vaccine strain 17D was provided by the Institut Pasteur de Dakar vaccine production unit (reference 1927). Briefly, the yellow fever vaccine is prepared using various seed strains that were ultimately derived from the 17D strain of yellow fever virus cultured in chicken embryonated eggs. The vaccine was obtained lyophilized. For viral stock preparation, each vial was reconstituted with 5ml of buffer (NaCl 0,9%) and amplified in C636 cells. The passage of the virus stock is 2.

A defined virus concentration of $10^3$ plaque forming units (pfu) per ml (dose test, DT) and diluted, heat-inactivated sera (10-fold in Leibovitz's medium containing 3% fetal calf serum, FCS, heated for 20 min at 60°C) were titrated in serial two-fold dilutions from 1:10 to 1:20480. 30 µl of each serum dilution and 30µl of virus were mixed and pre-incubated at 37°C for 1 hour. Calibration range was done with different doses of the challenge virus (DT, 50, 30, 10, 5 and 1% of the DT). Wells with cells alone were used as negative control and wells with undiluted virus as positive controls. In addition, a serum provided by NIBSC was used as international standard for the validation of the calibration range. After 1-hour incubation, $10^6$ cells/ml of PS cells in L15-3 % FCS were then added to each well containing the mixture serum/virus and all control wells and incubated for 4 hours at 37°C before overlaying with 0.6% carboxy-methyl-cellulose and 3% FCS in L-15 Leibovitz's media. Plates were sealed and incubated for 4 to 5 days at 37°C for the development of cytopathic effect. Wells were washed in phosphate buffer saline and stained in amino black solution (0.1%) for 30 minutes at room temperature. Plates were rinsed with water and allowed to dry then infectious plaques visualized.

The test was considered valid if: (1) there was no lysis plaques in the negative control wells; (2) total lysis in the undiluted virus wells; and, (3) linear lysis range in wells containing the different doses of the challenge virus (30-50 lysis plaques for the DT, 15-25 lysis plaques for the half dose and 3-5 lysis plaques for the 10% dose). Inhibition percentage of the DT for a serum was then obtained by comparing the results with the calibration range. Plaques were counted for each serum and antibody titer was determined as the antibody dilution that reduced observed lysis plaques by 90% and 50% compared to the number observed in the calibration range ($PRNT_{90}$ and $PRNT_{50}$).

### Seroconversion and geometric mean titers, per-protocol population

Table S1: Non-inferiority of seroconversion and GMT by PRNT90 in fractional vs. standard dose of yellow fever vaccine at Day 10, Day 28 and Day 365 post-vaccination, by vaccine manufacturer, in the per-protocol population

| Manufacturer | Time | Dosage | N | Seroconversion | | | GMT | |
|---|---|---|---|---|---|---|---|---|
| | | | | n | % (95% CI) | Difference, % (95% CI) | GMT (95% CI) | Ratio GMT (95% CI) |
| Bio-Manguinhos | D10 | Fractional | 110 | 8 | 7.3 (3.2, 13.8) | -2.13 (-9.74, 5.41) | 6.6 (5.9, 7.4) | 1.00 (0.85, 1.17) |
| | | Standard | 117 | 11 | 9.4 (4.8, 16.2) | | 6.6 (5.9, 7.4) | |
| | D28 | Fractional | 111 | 110 | 99.1 (95.1, 100.0) | 3.37 (-2.02, 8.08) | 379 (282, 509) | 0.89 (0.58, 1.34) |
| | | Standard | 117 | 112 | 95.7 (90.3, 98.6) | | 428 (318, 575) | |
| | D365 | Fractional | 110 | 102 | 92.7 (86.2, 96.8) | 6.24 (-2.14, 14.5) | 95.4 (73.3, 124) | 1.22 (0.84, 1.79) |
| | | Standard | 111 | 96 | 86.5 (78.7, 92.2) | | 78.0 (59.3, 103) | |
| Chumakov IPVE | D10 | Fractional | 111 | 8 | 7.2 (3.2, 13.7) | -10.34 (-19.01, -1.59) | 6.3 (5.7, 6.9) | 0.83 (0.71, 0.97) |
| | | Standard | 114 | 20 | 17.5 (11.1, 25.8) | | 7.5 (6.6, 8.5) | |
| | D28 | Fractional | 111 | 105 | 94.6 (88.6, 98.0) | -2.77 (-8.40, 3.36) | 561 (399,789) | 1.14 (0.74, 1.76) |
| | | Standard | 114 | 111 | 97.4 (92.5, 99.5) | | 493 (375, 648) | |
| | D365 | Fractional | 107 | 98 | 91.6 (84.6, 96.1) | -2.16 (-9.41, 5.35) | 125.9 (94.4, 168) | 1.40 (0.96, 2.03) |
| | | Standard | 112 | 105 | 93.8 (87.5, 97.5) | | 90.0 (70.9, 114) | |
| IPD | D10 | Fractional | 112 | 16 | 14.3 (8.4, 22.2) | -14.42 (-25.27, -3.85) | 7.2 (6.4, 8.2) | 0.74 (0.60, 0.92) |
| | | Standard | 108 | 31 | 28.7 (20.4, 38.2) | | 9.7 (8.2, 11.4) | |
| | D28 | Fractional | 112 | 111 | 99.1 (95.1, 100.0) | 3.65 (-2.05, 8.41) | 376 (294, 481) | 1.35 (0.92, 1.98) |
| | | Standard | 110 | 105 | 95.5 (89.7, 98.5) | | 279 (208, 374) | |
| | D365 | Fractional | 111 | 105 | 94.6 (88.6, 98.0) | 0.25 (-6.54, 6.88) | 106 (82.7, 136) | 1.40 (0.99, 1.98) |
| | | Standard | 106 | 100 | 94.3 (88.1, 97.9) | | 75.9 (59.3, 97.1) | |
| Sanofi Pasteur | D10 | Fractional | 113 | 14 | 12.4 (6.9, 19.9) | -18.24 (-28.96, -7.92) | 6.9 (6.2, 7.7) | 0.71 (0.59, 0.86) |
| | | Standard | 111 | 34 | 30.6 (22.2, 40.1) | | 9.7 (8.3, 11.3) | |
| | D28 | Fractional | 113 | 110 | 97.3 (92.4, 99.5) | -1.76 (-6.12, 3.15) | 498 (385, 644) | 1.21 (0.84, 1.76) |
| | | Standard | 112 | 111 | 99.1 (95.1, 100.0) | | 410 (312, 538) | |
| | D365 | Fractional | 109 | 104 | 95.4 (89.6, 98.5) | -0.92 (-6.91, 5.21) | 95.6 (76.7, 119) | 0.82 (0.58, 1.14) |
| | | Standard | 109 | 105 | 96.3 (90.9, 99.0) | | 117 (90.3, 152) | |

### Seroconversion, additional analysis

Table S2: Non-inferiority of seroconversion rate by $PRNT_{50}$ in fractional vs. standard dose of yellow fever vaccine at Day 28 in intent-to-treat population, by vaccine manufacturer

| Manufacturer | Fractional dose | | Standard dose | | Difference (Fractional - Standard) |
|---|---|---|---|---|---|
| | No. SC | % SC (95% CI) | No. SC | % SC (95% CI) | (95% CI) |
| Bio-Manguinhos | 117/117 | 100.0 (96.9, 100.0) | 116/118 | 98.3 (94.0, 99.8) | 1.69 (-2.58, 5.10) |
| Chumakov IPVE | 115/118 | 97.5 (92.7, 99.5) | 118/118 | 100.0 (96.9, 100.0) | -2.54 (-6.11, 2.12) |
| IPD | 119/119 | 100.0 (96.9, 100.0) | 116/119 | 97.5 (92.8, 99.5) | 2.52 (-2.11, 6.06) |
| Sanofi Pasteur | 120/120 | 100.0 (97.0, 100.0) | 120/120 | 100.0 (97.0, 100.0) | 0.0 (-3.10, 3.10) |

Table S3: Non-inferiority of seroconversion rate by $PRNT_{50}$ in fractional vs. standard dose of yellow fever vaccine at Day 28 in intent-to-treat population with baseline seropositivity to yellow fever, by vaccine manufacturer

| Manufacturer | Fractional dose | | Standard dose | | Difference (Fractional - Standard) |
|---|---|---|---|---|---|
| | No. SC | % SC (95% CI) | No. SC | % SC (95% CI) | (95% CI) |
| Bio-Manguinhos | 6/6 | 100.0 (54.1, 100.0) | 1/1 | 100.0 (2.5, 100.0) | 0.0 (-79.3, 39.0) |
| Chumakov IPVE | 5/7 | 71.4 (29.0, 96.3) | 4/4 | 100.0 (39.8, 100.0) | -28.57 (-81.6, 7.0) |
| IPD | 7/7 | 100.0 (59.0, 100.0) | 8/9 | 88.9 (51.8, 99.7) | 11.1 (-21.3, 47.7) |
| Sanofi Pasteur | 7/7 | 100.0 (59.0, 100.0) | 8/8 | 100.0 (63.1, 100.0) | 0.0 (-32.4, 35.4) |

### Geometric mean titers, additional analysis

Table S4: Geometric mean titer by $PRNT_{50}$ in fractional vs. standard dose of yellow fever vaccine at Day 28 in intent-to-treat population, by vaccine manufacturer

| Manufacturer | Fractional dose | | | Standard dose | | | Difference - log GMT (Fractional – Standard) | Ratio - GMT (Fractional/ Standard) |
|---|---|---|---|---|---|---|---|---|
| | N | GMT | 95% CI | N | GMT | 95% CI | (95% CI) | (95% CI) |
| Bio-Manguinhos | 117 | 3718.2 | 2676.0, 5166.3 | 118 | 4119.8 | 2895.4, 5862.1 | -0.045 (-0.253, 0.164) | 0.90 (0.56, 1.46) |
| Chumakov IPVE | 118 | 5826.3 | 4152.9, 8174.1 | 118 | 5493.9 | 4045.0, 7461.9 | 0.026 (-0.172, 0.223) | 1.06 (0.67, 1.67) |
| IPD | 119 | 4224.7 | 3170.7, 5629.0 | 119 | 2530.4 | 1787.6, 3581.8 | 0.223 (0.028, 0.417) | 1.67 (1.07, 2.61) |
| Sanofi Pasteur | 120 | 5713.9 | 4290.7, 7609.1 | 120 | 4280.6 | 3105.3, 5900.7 | 0.125 (-0.060, 0.311) | 1.33 (0.87, 2.05) |

Table S5: Geometric mean titer by $PRNT_{50}$ in fractional vs. standard dose of yellow fever vaccine at Day 28 in intent-to-treat population with baseline seropositivity to yellow fever, by vaccine manufacturer

| Manufacturer | Fractional dose | | | Standard dose | | | Difference - log GMT (Fractional – Standard) | Ratio - GMT (Fractional/ Standard) |
|---|---|---|---|---|---|---|---|---|
| | N | GMT | 95% CI | N | GMT | 95% CI | (95% CI) | (95% CI) |
| Bio-Manguinhos | 6 | 1280.0 | 215.5, 7603.8 | 1 | 20480 | -- | -1.204 (-3.251, 0.843) | 0.063 (0.001, 6.970) |
| Chumakov IPVE | 7 | 5120.0 | 555.7, 47173.5 | 4 | 1076.3 | 22.7, 51106.1 | 0.667 (-0.914, 2.268) | 4.76 (0.12, 185.45) |
| IPD | 7 | 3445.5 | 721.1, 16462.5 | 9 | 2031.9 | 522.3, 7903.7 | 0.229 (-0.584, 1.043) | 1.70 (0.26, 11.04) |
| Sanofi Pasteur | 7 | 9274.6 | 3628.7, 23705.0 | 8 | 4695.1 | 1447.1, 15232.8 | 0.296 (-0.296, 0.887) | 1.98 (0.51, 7.71) |

Figure S1: Reverse cumulative distributions of geometric mean titers by $PRNT_{50}$ at Day 28 for per-protocol population, by vaccine manufacturer and by vaccine dose

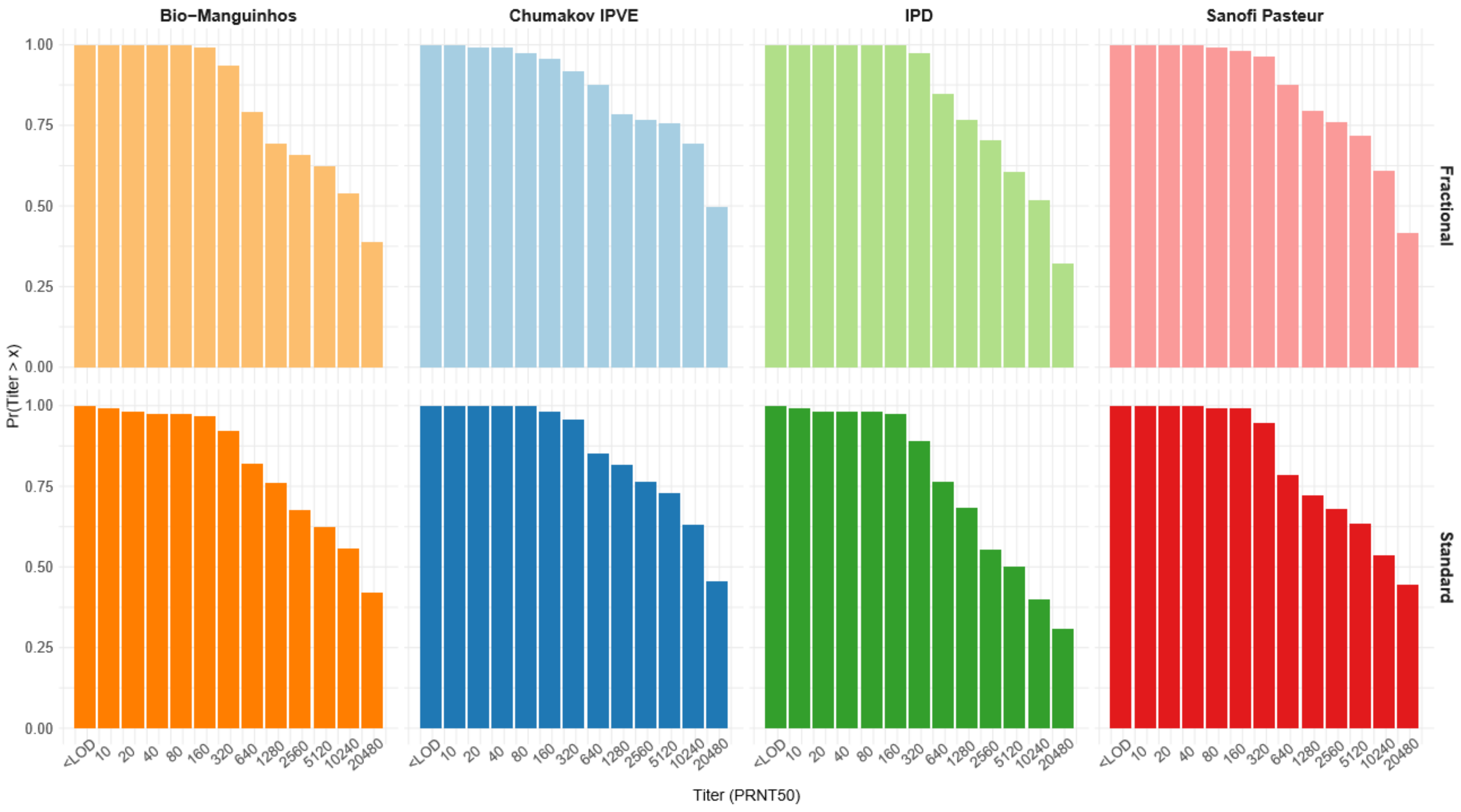

Figure S2: Reverse cumulative distributions of geometric mean titers by PRNT50 at Day 10 for per-protocol population, by vaccine manufacturer and by vaccine dose

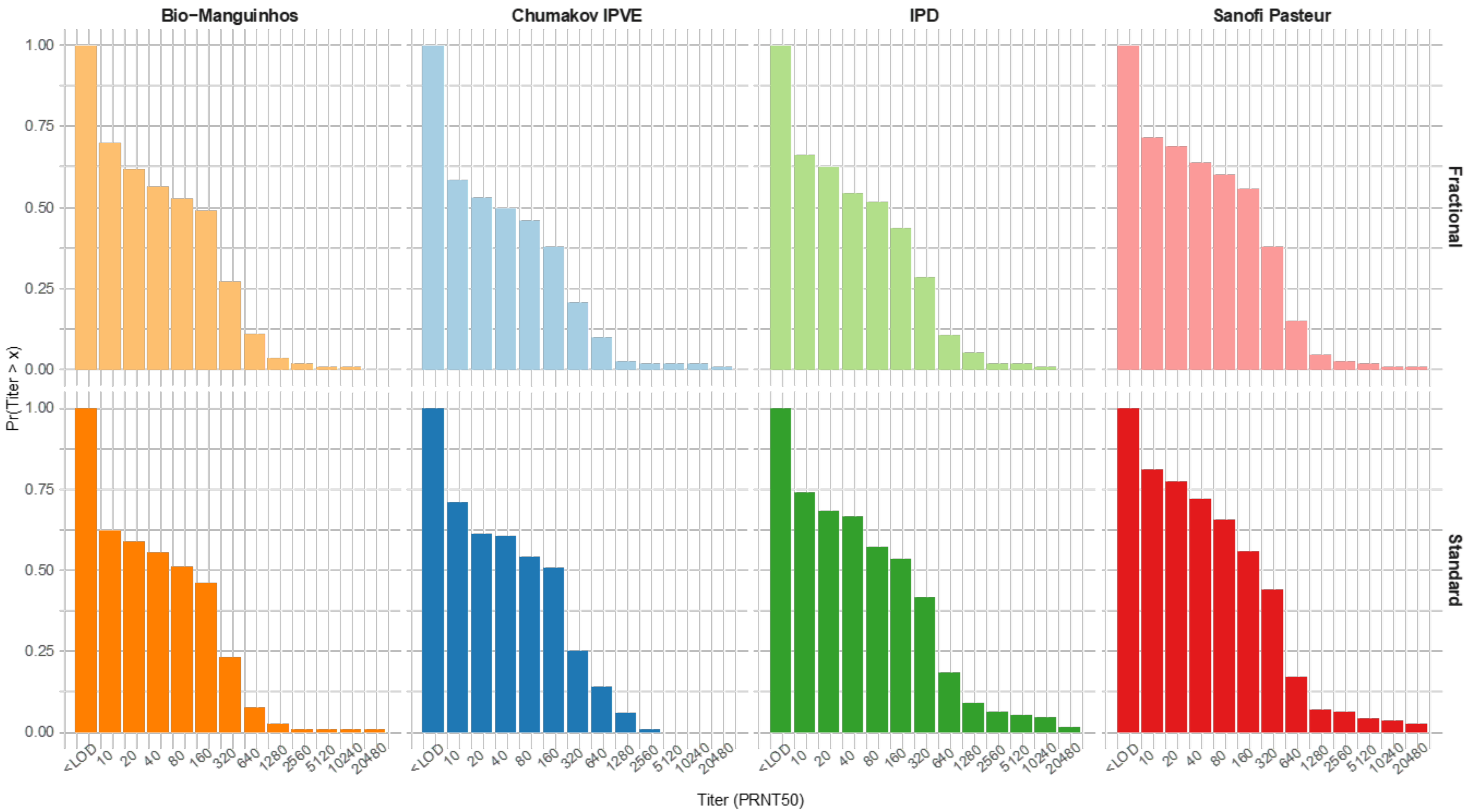

Figure S3: Reverse cumulative distributions of geometric mean titers by $PRNT_{50}$ at Day 28 for intent-to-treat population with baseline seropositivity to yellow fever, by vaccine manufacturer and by vaccine dose

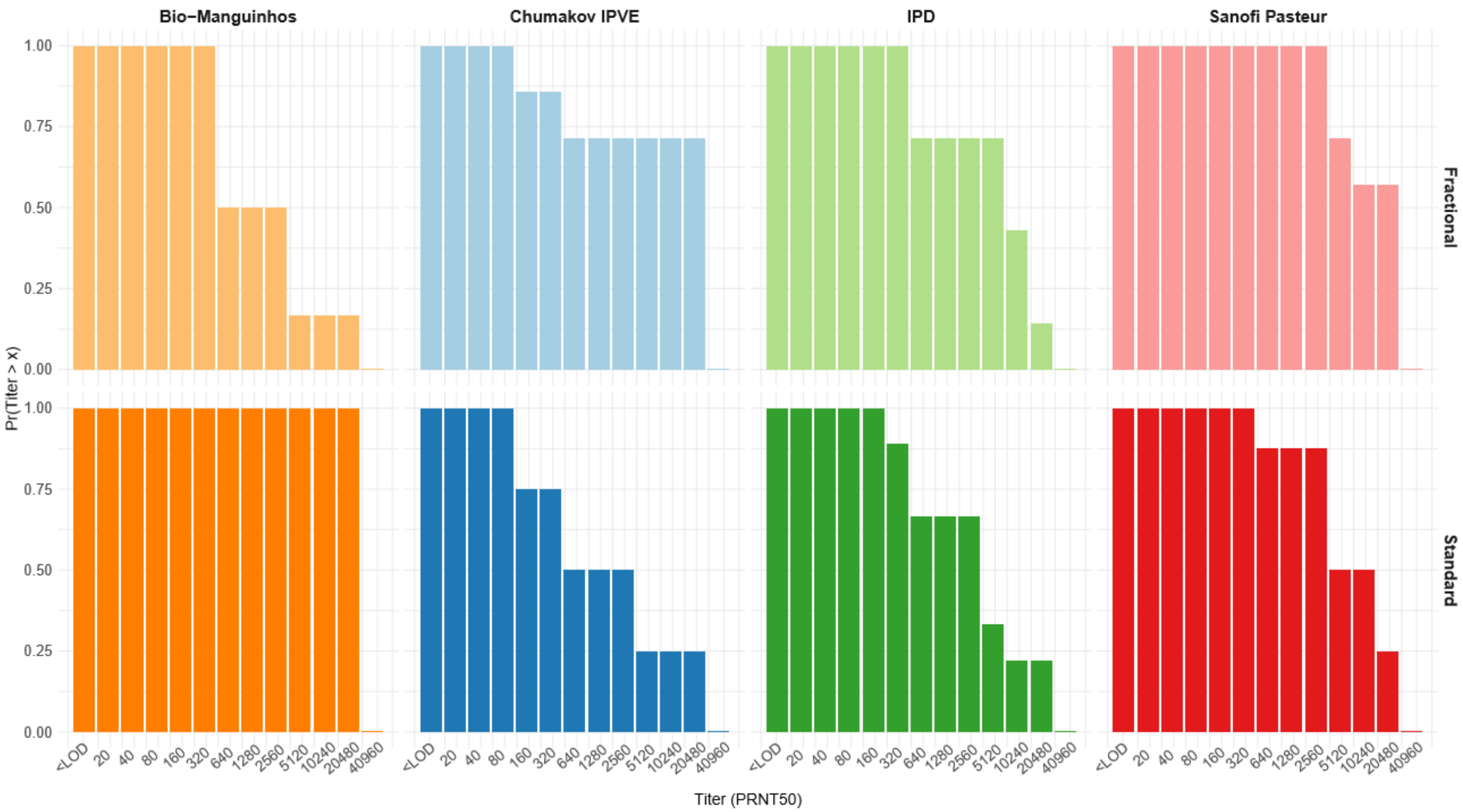

Figure S4: Reverse cumulative distributions of geometric mean titers by $PRNT_{50}$ at Day 10 for intent-to-treat population, by vaccine manufacturer and by vaccine dose

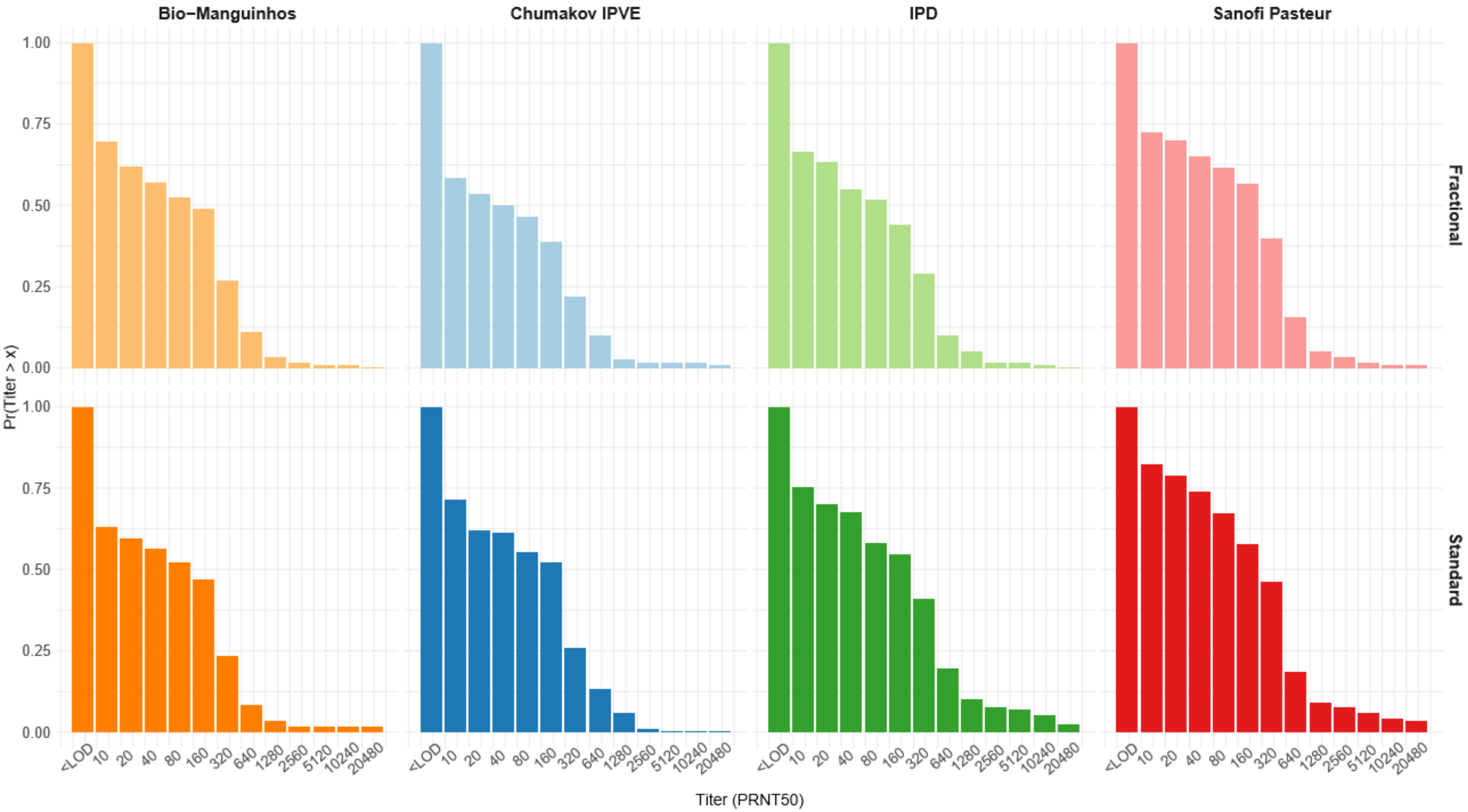

Figure S5: Reverse cumulative distributions of geometric mean titers by $PRNT_{50}$ at Day 365 for intent-to-treat population, by vaccine manufacturer and by vaccine dose

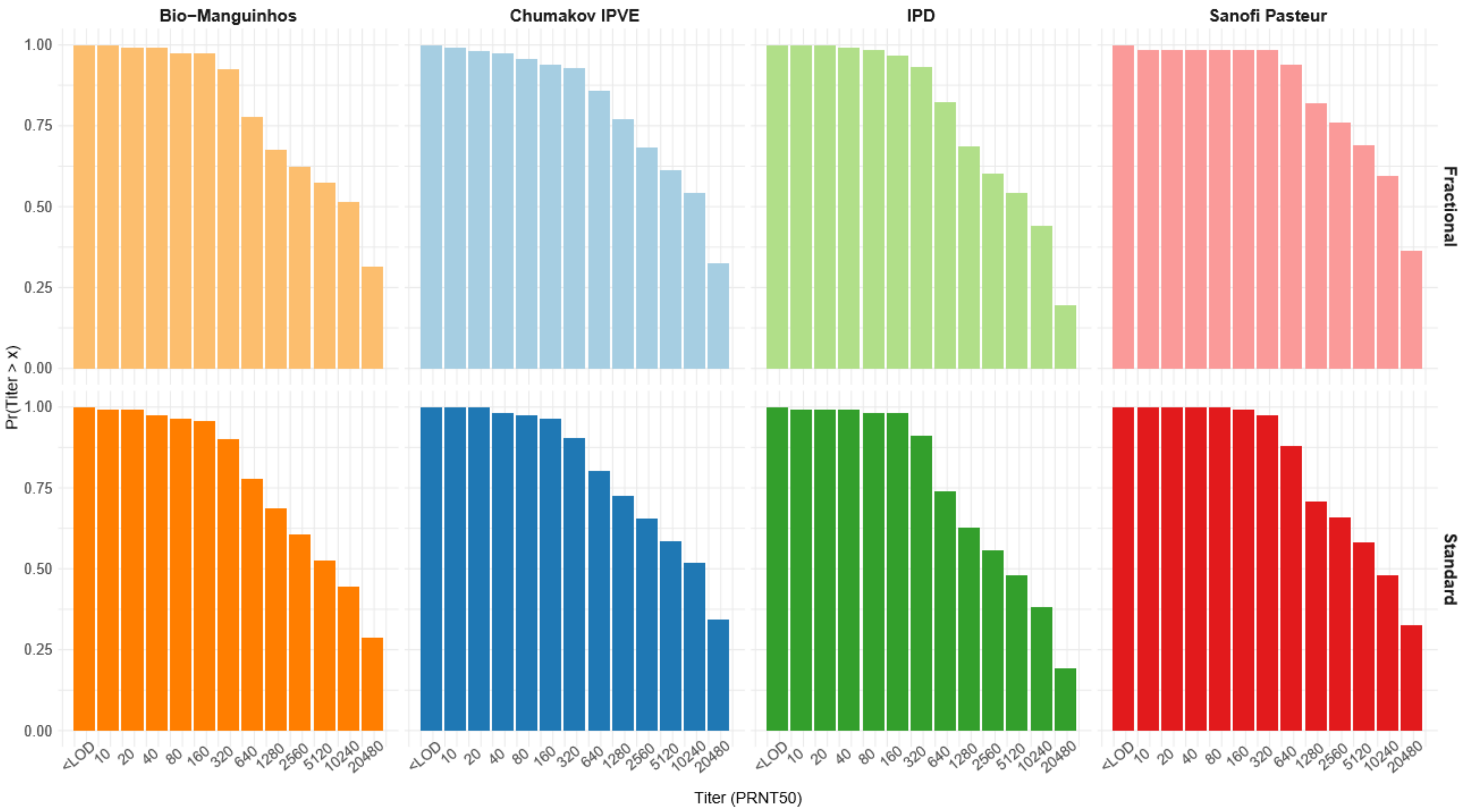

Figure S6: Geometric mean titer by $PRNT_{50}$ at various timepoints post-vaccination for ITT population, by vaccine manufacturer and by vaccine dose.

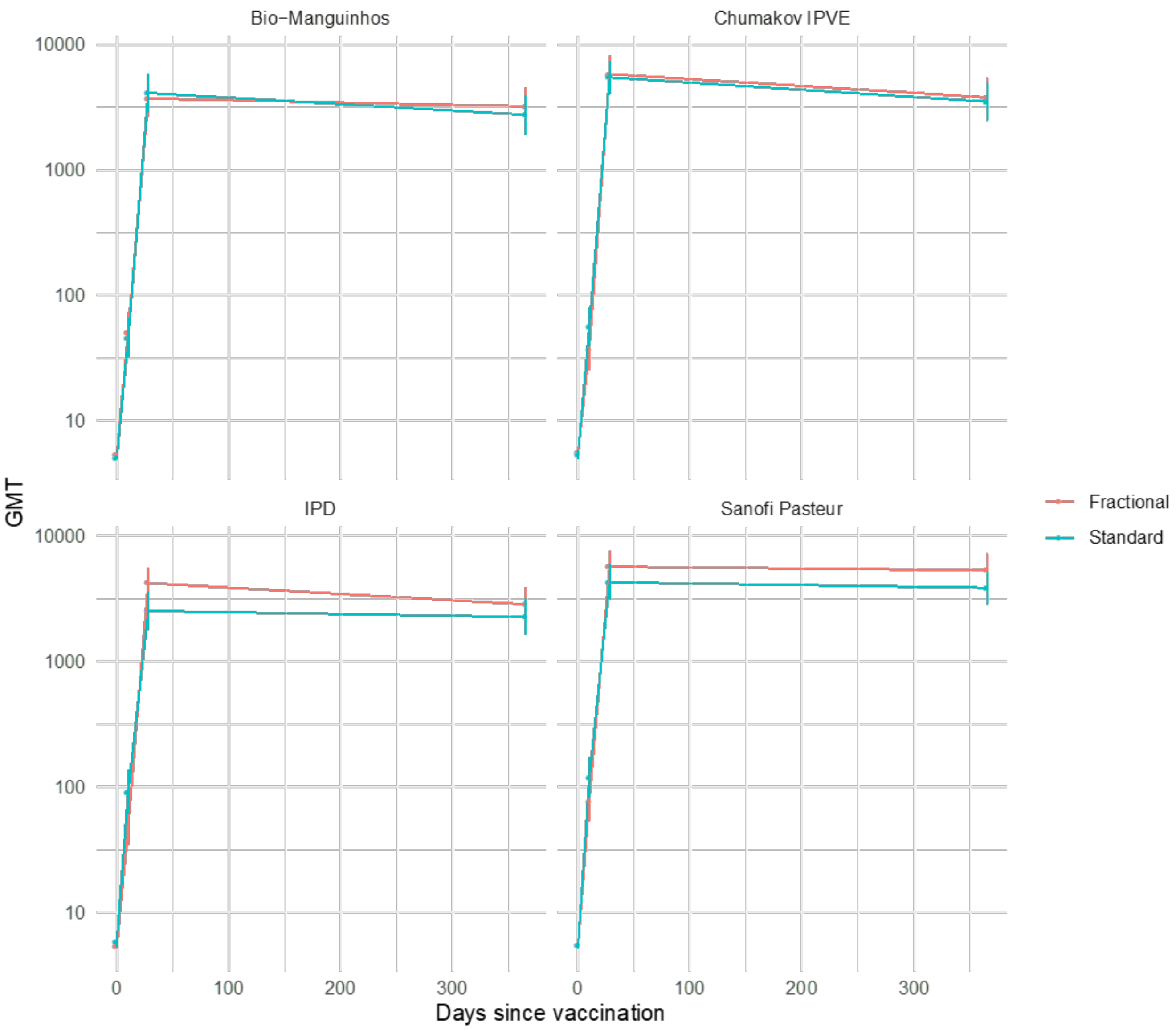

Note: vertical bars represent 95% confidence intervals.

Geometric mean fold increase

Table S6: Geometric mean fold increase by $PRNT_{50}$ in fractional vs. standard dose of yellow fever vaccine at Day 28 in per-protocol population, by vaccine manufacturer

| Manufacturer | Fractional dose | | | Standard dose | | | Difference - log GMFI (Fractional – Standard) | Ratio - GMFI (Fractional/ Standard) |
|---|---|---|---|---|---|---|---|---|
| | N | GM FI | 95% CI | N | GMFI | 95% CI | (95% CI) | (95% CI) |
| Bio-Manguinhos | 111 | 788 | 562, 1103 | 117 | 813 | 570, 1159 | -0.014 (-0.225, 0.198) | 0.97 (0.60, 1.58) |
| Chumakov IPVE | 111 | 1175 | 832, 1658 | 114 | 1163 | 859.3, 1575 | 0.004 (-0.194, 0.202) | 1.01 (0.64, 1.59) |
| IPD | 112 | 856 | 636, 1151 | 110 | 515 | 358, 742 | 0.220 (0.017, 0.423) | 1.66 (1.04, 2.65) |
| Sanofi Pasteur | 113 | 1109 | 821, 1498 | 112 | 850 | 607, 1192 | 0.115 (-0.080, 0.311) | 1.30 (0.83, 2.04) |

Table S7: Geometric mean fold increase by $PRNT_{90}$ in fractional vs. standard dose of yellow fever vaccine at Day 28 in per-protocol population, by vaccine manufacturer

| Manufacturer | Fractional dose | | | Standard dose | | | Difference - log GMFI (Fractional – Standard) | Ratio - GMFI (Fractional/ Standard) |
|---|---|---|---|---|---|---|---|---|
| | N | GMFI | 95% CI | N | GMFI | 95% CI | (95% CI) | (95% CI) |
| Bio-Manguinhos | 111 | 75.8 | 56.4, 101.8 | 117 | 85.6 | 63.6, 115.1 | -0.053 (-0.234, 0.128) | 0.89 (0.58, 1.34) |
| Chumakov IPVE | 111 | 112.3 | 79.9, 157.8 | 114 | 98.6 | 75.0, 129.5 | 0.057 (-0.132, 0.245) | 1.14 (0.74, 1.76) |
| IPD | 112 | 75.2 | 58.8, 96.1 | 110 | 55.7 | 41.5, 74.8 | 0.130 (-0.036, 0.296) | 1.35 (0.92, 1.98) |
| Sanofi Pasteur | 113 | 99.5 | 76.9, 128.8 | 112 | 82.0 | 62.4, 107.7 | 0.084 (-0.078, 0.246) | 1.21 (0.84, 1.76) |

Table S8: Geometric mean fold increase by $PRNT_{50}$ in fractional vs. standard dose of yellow fever vaccine at Day 28 in intent-to-treat population, by vaccine manufacturer

| Manufacturer | Fractional dose | | | Standard dose | | | Difference - log GMFI (Fractional – Standard) | Ratio - GMFI (Fractional/ Standard) |
|---|---|---|---|---|---|---|---|---|
| | N | GMFI | 95% CI | N | GMFI | 95% CI | (95% CI) | (95% CI) |
| Bio-Manguinhos | 117 | 696.7 | 495.7, 979.4 | 118 | 809.6 | 569.5, 1150.8 | -0.065 (-0.277, 0.146) | 0.86 (0.53, 1.40) |
| Chumakov IPVE | 118 | 1036.1 | 716.6, 1498.1 | 118 | 1018.0 | 732.6, 1414.6 | 0.008 (-0.206, 0.221) | 1.02 (0.62, 1.66) |
| IPD | 119 | 787.9 | 588.0, 1055.7 | 119 | 432.4 | 299.8, 623.7 | 0.261 (0.058, 0.463) | 1.82 (1.14, 2.91) |
| Sanofi Pasteur | 120 | 1047.9 | 780.5, 1406.9 | 120 | 789.6 | 571.0, 1091.8 | 0.123 (-0.066, 0.312) | 1.33 (0.86, 2.05) |

Table S9: Geometric mean fold increase in by $PRNT_{50}$ fractional vs. standard dose of yellow fever vaccine at Day 28 in intent-to-treat population with baseline seropositivity to yellow fever, by vaccine manufacturer

| Manufacturer | Fractional dose | | | Standard dose | | | Difference - log GMFI (Fractional – Standard) | Ratio - GMFI (Fractional/ Standard) |
|---|---|---|---|---|---|---|---|---|
| | N | GMFI | 95% CI | N | GMFI | 95% CI | (95% CI) | (95% CI) |
| Chumakov IPVE | 7 | 141.3 | 4.8, 4153.2 | 4 | 22.6 | 0.4, 1334.9 | 0.796 (-1.077, 2.668) | 6.25 (0.08, 465.7) |
| IPD | 7 | 210.0 | 46.2, 953.6 | 9 | 50.8 | 11.8, 218.5 | 0.616 (-0.209, 1.442) | 4.13 (0.62, 27.6) |
| Sanofi Pasteur | 7 | 420.0 | 81.2, 2172.5 | 8 | 279.2 | 93.6, 832.3 | 0.177 (-0.602, 0.957) | 1.50 (0.25, 9.06) |
| Bio-Manguinhos | 6 | 71.8 | 11.1, 463.3 | 1 | 512 | -- | -0.853 (-2.995, 1.289) | 0.14 (0.001, 19.5) |

Table S10: Geometric mean fold increase in by $PRNT_{50}$ fractional vs. standard dose of yellow fever vaccine at Day 10, in intent-to-treat population, by vaccine manufacturer

| Manufacturer | Fractional dose | | | Standard dose | | | Difference - log GMFI (Fractional – Standard) | Ratio - GMFI (Fractional/ Standard) |
|---|---|---|---|---|---|---|---|---|
| | N | GMFI | 95% CI | N | GMFI | 95% CI | (95% CI) | (95% CI) |
| Bio-Manguinhos | 116 | 9.7 | 6.8, 14.0 | 119 | 9.0 | 6.3, 12.9 | 0.033 (-0.187, 0.252) | 1.08 (0.65, 1.79) |
| Chumakov IPVE | 118 | 6.6 | 4.5, 9.5 | 119 | 10.4 | 7.3, 14.9 | -0.201 (-0.424, 0.022) | 0.63 (0.38, 1.05) |
| IPD | 120 | 9.1 | 6.3, 13.2 | 117 | 15.5 | 10.3, 23.3 | -0.230 (-0.468, 0.007) | 0.59 (0.34, 1.02) |
| Sanofi Pasteur | 120 | 14.0 | 9.7, 20.2 | 119 | 21.7 | 14.9, 31.4 | -0.189 (-0.414, 0.036) | 0.65 (0.39, 1.09) |

Table S11: Geometric mean fold increase in by $PRNT_{50}$ fractional vs. standard dose of yellow fever vaccine at Day 365, in intent-to-treat population, by vaccine manufacturer

| Manufacturer | Fractional dose | | | Standard dose | | | Difference - log GMFI (Fractional – Standard) | Ratio - GMFI (Fractional/ Standard) |
|---|---|---|---|---|---|---|---|---|
| | N | GMFI | 95% CI | N | GMFI | 95% CI | (95% CI) | (95% CI) |
| Bio-Manguinhos | 117 | 604 | 423, 864 | 112 | 541 | 376, 779 | 0.048 (-0.172, 0.268) | 1.12 (0.67, 1.85) |
| Chumakov IPVE | 114 | 673 | 456, 995 | 116 | 650 | 449, 941 | 0.015 (-0.217, 0.247) | 1.04 (0.61, 1.77) |
| IPD | 118 | 533 | 386, 738 | 115 | 386 | 272, 547 | 0.141 (-0.065, 0.347) | 1.38 (0.86, 2.22) |
| Sanofi Pasteur | 116 | 976 | 722, 1320 | 117 | 713 | 525, 969 | 0.136 (-0.049, 0.322) | 1.37 (0.89, 2.10) |

### Safety Analysis

#### Adverse events

Table S12: Summary of adverse events up to Day 28 post-vaccination by study arm, and vaccine manufacturer, safety population

| Manufacturer | Fractional Dose | | Standard Dose | | P value |
|---|---|---|---|---|---|
| | N | No. with ≥1 AE (%) | N | No. with ≥1 AE (%) | |
| Bio-Manguinhos | 120 | 58 (48.3) | 120 | 69 (57.5) | 0.20 |
| Chumakov IPVE | 119 | 69 (58.0) | 120 | 72 (60.0) | 0.85 |
| IPD | 120 | 76 (63.3) | 120 | 72 (60.0) | 0.69 |
| Sanofi Pasteur | 120 | 60 (50.0) | 120 | 70 (53.8) | 0.24 |

Table S13: Adverse events up to Day 28 post-vaccination by MedDRA coding (v20.0), study arm and vaccine manufacturer, safety population

| System Organ Class and Preferred Term | Bio-Manguinhos | | Chumakov IPVE | | IPD | | Sanofi Pasteur | |
|---|---|---|---|---|---|---|---|---|
| n (%) with ≥ 1 AE | Frac. N=120 | Standard N=120 | Frac. N=119 | Standard N=120 | Frac. N=120 | Standard N=120 | Frac. N=120 | Standard N=120 |
| **Blood and lymphatic system disorders** | 0 (0) | 0 (0) | 0 (0) | 0 (0) | 0 (0) | 0 (0) | 1 (0.8) | 1 (0.8) |
| Anemia | 0 (0) | 0 (0) | 0 (0) | 0 (0) | 0 (0) | 0 (0) | 1 (0.8) | 0 (0) |
| Lymphadenopathy | 0 (0) | 0 (0) | 0 (0) | 0 (0) | 0 (0) | 0 (0) | 0 (0) | 1 (0.8) |
| **Cardiac disorders** | 0 (0) | 0 (0) | 1 (0.8) | 0 (0) | 0 (0) | 0 (0) | 0 (0) | 0 (0) |
| Palpitations | 0 (0) | 0 (0) | 1 (0.8) | 0 (0) | 0 (0) | 0 (0) | 0 (0) | 0 (0) |
| **Ear and labyrinth disorders** | 1 (0.8) | 0 (0) | 0 (0) | 0 (0) | 1 (0.8) | 1 (0.8) | 0 (0) | 1 (0.8) |
| Allergic Otitis Externa | 0 (0) | 0 (0) | 0 (0) | 0 (0) | 0 (0) | 0 (0) | 0 (0) | 1 (0.8) |
| Ear Pain | 0 (0) | 0 (0) | 0 (0) | 0 (0) | 0 (0) | 1 (0.8) | 0 (0) | 0 (0) |
| Sudden Hearing Loss | 1 (0.8) | 0 (0) | 0 (0) | 0 (0) | 1 (0.8) | 0 (0) | 0 (0) | 0 (0) |
| **Eye disorders** | 0 (0) | 0 (0) | 2 (1.7) | 1 (0.8) | 2 (1.7) | 0 (0) | 0 (0) | 0 (0) |
| Conjunctivitis Allergic | 0 (0) | 0 (0) | 2 (1.7) | 0 (0) | 2 (1.7) | 0 (0) | 0 (0) | 0 (0) |
| Eye Pruritus | 0 (0) | 0 (0) | 0 (0) | 1 (0.8) | 0 (0) | 0 (0) | 0 (0) | 0 (0) |
| **Gastrointestinal disorders** | 8 (6.7) | 14 (11.7) | 15 (12.6) | 15 (12.5) | 13 (10.8) | 12 (10.0) | 11 (9.2) | 13 (10.8) |
| Abdominal discomfort | 0 (0) | 0 (0) | 1 (0.8) | 0 (0) | 0 (0) | 0 (0) | 0 (0) | 0 (0) |
| Abdominal distension | 0 (0) | 0 (0) | 0 (0) | 0 (0) | 0 (0) | 0 (0) | 1 (0.8) | 0 (0) |
| Abdominal pain | 4 (3.3) | 3 (2.5) | 3 (2.5) | 8 (6.7) | 9 (7.5) | 1 (0.8) | 6 (5.0) | 3 (2.5) |
| Diarrhea | 2 (1.7) | 2 (1.7) | 4 (3.4) | 2 (1.7) | 0 (0) | 5 (4.2) | 0 (0) | 0 (0) |
| Dyspepsia | 0 (0) | 0 (0) | 0 (0) | 0 (0) | 0 (0) | 1 (0.8) | 0 (0) | 0 (0) |
| Epigastric discomfort | 0 (0) | 0 (0) | 1 (0.8) | 0 (0) | 0 (0) | 0 (0) | 0 (0) | 0 (0) |
| Food poisoning | 0 (0) | 0 (0) | 0 (0) | 0 (0) | 0 (0) | 0 (0) | 0 (0) | 1 (0.8) |
| Gastritis | 0 (0) | 1 (0.8) | 2 (1.7) | 1 (0.8) | 2 (1.7) | 0 (0) | 1 (0.8) | 2 (1.7) |
| Gastrointestinal disorder | 2 (1.7) | 0 (0) | 0 (0) | 1 (0.8) | 1 (0.8) | 2 (1.7) | 1 (0.8) | 0 (0) |
| Gastroesophageal reflux disease | 0 (0) | 1 (0.8) | 0 (0) | 0 (0) | 0 (0) | 0 (0) | 0 (0) | 0 (0) |
| Nausea | 1 (0.8) | 6 (5.0) | 3 (2.5) | 2 (1.7) | 2 (1.7) | 3 (2.5) | 2 (1.7) | 7 (5.8) |
| Peptic ulcer | 0 (0) | 0 (0) | 1 (0.8) | 0 (0) | 0 (0) | 0 (0) | 0 (0) | 1 (0.8) |
| Stomatitis | 0 (0) | 0 (0) | 0 (0) | 1 (0.8) | 0 (0) | 0 (0) | 0 (0) | 0 (0) |
| Toothache | 0 (0) | 1 (0.8) | 0 (0) | 0 (0) | 0 (0) | 0 (0) | 0 (0) | 0 (0) |
| Vomiting | 0 (0) | 0 (0) | 1 (0.8) | 0 (0) | 0 (0) | 0 (0) | 1 (0.8) | 0 (0) |

| | | | | | | | | |
|---|---|---|---|---|---|---|---|---|
| **General disorders and administration site conditions** | 19 (15.8) | 25 (20.8) | 29 (24.4) | 21 (17.5) | 29 (24.2) | 25 (20.8) | 30 (25.0) | 25 (20.8) |
| Chest pain | 1 (0.8) | 0 (0) | 0 (0) | 1 (0.8) | 0 (0) | 0 (0) | 0 (0) | 0 (0) |
| Fatigue | 8 (6.7) | 15 (12.5) | 18 (15.1) | 12 (10.0) | 19 (15.8) | 21 (17.5) | 21 (17.5) | 17 (14.2) |
| Injection site discomfort | 0 (0) | 0 (0) | 0 (0) | 1 (0.8) | 0 (0) | 0 (0) | 0 (0) | 0 (0) |
| Injection site hypoesthesia | 0 (0) | 0 (0) | 0 (0) | 0 (0) | 0 (0) | 0 (0) | 1 (0.8) | 0 (0) |
| Injection site reaction | 1 (0.8) | 0 (0) | 0 (0) | 0 (0) | 0 (0) | 0 (0) | 0 (0) | 0 (0) |
| Local reaction | 1 (0.8) | 1 (0.8) | 1 (0.8) | 1 (0.8) | 0 (0) | 0 (0) | 0 (0) | 1 (0.8) |
| Malaise | 0 (0) | 0 (0) | 0 (0) | 1 (0.8) | 0 (0) | 0 (0) | 0 (0) | 0 (0) |
| Pyrexia | 9 (7.5) | 11 (9.2) | 12 (10.1) | 9 (7.5) | 12 (10.0) | 9 (7.5) | 10 (8.3) | 14 (11.7) |
| Vaccination site discomfort | 0 (0) | 1 (0.8) | 0 (0) | 1 (0.8) | 1 (0.8) | 0 (0) | 0 (0) | 0 (0) |
| Vaccination site irritation | 0 (0) | 1 (0.8) | 0 (0) | 0 (0) | 0 (0) | 0 (0) | 0 (0) | 0 (0) |
| Vaccination site pain | 1 (0.8) | 0 (0) | 0 (0) | 0 (0) | 0 (0) | 0 (0) | 1 (0.8) | 0 (0) |
| Vaccination site paresthesia | 0 (0) | 0 (0) | 0 (0) | 0 (0) | 0 (0) | 0 (0) | 1 (0.8) | 0 (0) |
| **Infections and infestations** | 16 (13.3) | 19 (15.8) | 14 (11.8) | 15 (12.5) | 20 (16.7) | 18 (15.0) | 9 (7.5) | 15 (12.5) |
| Abscess | 0 (0) | 1 (0.8) | 0 (0) | 0 (0) | 0 (0) | 0 (0) | 0 (0) | 1 (0.8) |
| Acrodermatitis | 0 (0) | 0 (0) | 1 (0.8) | 0 (0) | 0 (0) | 0 (0) | 0 (0) | 0 (0) |
| Bacterial vaginosis | 1 (0.8) | 0 (0) | 0 (0) | 0 (0) | 1 (0.8) | 1 (0.8) | 1 (0.8) | 0 (0) |
| Dysentery | 0 (0) | 1 (0.8) | 0 (0) | 0 (0) | 0 (0) | 0 (0) | 0 (0) | 0 (0) |
| Eye infection, bacterial | 0 (0) | 0 (0) | 0 (0) | 0 (0) | 0 (0) | 0 (0) | 1 (0.8) | 0 (0) |
| Folliculitis | 0 (0) | 0 (0) | 0 (0) | 1 (0.8) | 0 (0) | 0 (0) | 0 (0) | 0 (0) |
| Fungal skin infection | 1 (0.8) | 0 (0) | 0 (0) | 1 (0.8) | 0 (0) | 0 (0) | 0 (0) | 0 (0) |
| Gastroenteritis | 0 (0) | 0 (0) | 0 (0) | 2 (1.7) | 0 (0) | 2 (1.7) | 1 (0.8) | 1 (0.8) |
| Genitourinary tract infection | 0 (0) | 0 (0) | 0 (0) | 1 (0.8) | 0 (0) | 0 (0) | 0 (0) | 0 (0) |
| Lower respiratory tract infection | 0 (0) | 0 (0) | 0 (0) | 1 (0.8) | 1 (0.8) | 0 (0) | 0 (0) | 0 (0) |
| Mastitis | 0 (0) | 0 (0) | 0 (0) | 0 (0) | 0 (0) | 1 (0.8) | 0 (0) | 1 (0.8) |
| Orchitis | 0 (0) | 0 (0) | 0 (0) | 1 (0.8) | 0 (0) | 0 (0) | 0 (0) | 0 (0) |
| Pelvic inflammatory disease | 0 (0) | 2 (1.7) | 1 (0.8) | 1 (0.8) | 0 (0) | 0 (0) | 0 (0) | 1 (0.8) |
| Pharyngitis | 0 (0) | 1 (0.8) | 1 (0.8) | 0 (0) | 2 (1.7) | 0 (0) | 1 (0.8) | 1 (0.8) |
| Respiratory tract infection | 3 (2.5) | 2 (1.7) | 4 (3.4) | 6 (5.0) | 6 (5.0) | 2 (1.7) | 1 (0.8) | 3 (2.5) |
| Respiratory tract infection, viral | 0 (0) | 0 (0) | 1 (0.8) | 0 (0) | 0 (0) | 0 (0) | 0 (0) | 0 (0) |
| Rhinitis | 0 (0) | 4 (3.3) | 1 (0.8) | 3 (2.5) | 3 (2.5) | 2 (1.7) | 0 (0) | 2 (1.7) |
| Skin bacterial infection | 0 (0) | 0 (0) | 1 (0.8) | 0 (0) | 0 (0) | 0 (0) | 0 (0) | 0 (0) |
| Tinea capitis | 0 (0) | 0 (0) | 1 (0.8) | 0 (0) | 0 (0) | 1 (0.8) | 0 (0) | 0 (0) |
| Tonsillitis | 1 (0.8) | 1 (0.8) | 1 (0.8) | 0 (0) | 1 (0.8) | 2 (1.7) | 2 (1.7) | 1 (0.8) |
| Upper respiratory tract infection | 4 (3.3) | 3 (2.5) | 0 (0) | 0 (0) | 4 (3.3) | 5 (4.2) | 1 (0.8) | 0 (0) |
| Urinary tract infection | 5 (4.2) | 2 (1.7) | 3 (2.5) | 0 (0) | 2 (1.7) | 2 (1.7) | 1 (0.8) | 1 (0.8) |
| Viral rhinitis | 1 (0.8) | 0 (0) | 1 (0.8) | 0 (0) | 0 (0) | 0 (0) | 0 (0) | 1 (0.8) |
| Viral upper respiratory tract infection | 0 (0) | 0 (0) | 0 (0) | 0 (0) | 1 (0.8) | 0 (0) | 0 (0) | 0 (0) |
| Vulvovaginal candidiasis | 0 (0) | 3 (2.5) | 0 (0) | 0 (0) | 0 (0) | 2 (1.7) | 0 (0) | 3 (2.5) |
| Wound sepsis | 0 (0) | 0 (0) | 0 (0) | 0 (0) | 0 (0) | 1 (0.8) | 1 (0.8) | 0 (0) |
| **Injury, poisoning, and procedural complaints** | 0 (0) | 0 (0) | 0 (0) | 0 (0) | 0 (0) | 0 (0) | 0 (0) | 2 (1.7) |
| Injury | 0 (0) | 0 (0) | 0 (0) | 0 (0) | 0 (0) | 0 (0) | 0 (0) | 1 (0.8) |

| | | | | | | | | |
|---|---|---|---|---|---|---|---|---|
| Road traffic accident | 0 (0) | 0 (0) | 0 (0) | 0 (0) | 0 (0) | 0 (0) | 0 (0) | 1 (0.8) |
| **Metabolism and nutrition disorders** | 1 (0.8) | 0 (0) | 0 (0) | 0 (0) | 0 (0) | 1 (0.8) | 0 (0) | 3 (2.5) |
| Abnormal loss of weight | 1 (0.8) | 0 (0) | 0 (0) | 0 (0) | 0 (0) | 0 (0) | 0 (0) | 0 (0) |
| Decreased appetite | 0 (0) | 0 (0) | 0 (0) | 0 (0) | 0 (0) | 1 (0.8) | 0 (0) | 3 (2.5) |
| **Musculoskeletal and connective tissue disorders** | 20 (16.7) | 22 (18.3) | 20 (16.8) | 19 (15.8) | 21 (17.5) | 26 (21.7) | 19 (15.8) | 19 (15.8) |
| Arthralgia | 4 (3.3) | 3 (2.5) | 4 (3.4) | 2 (1.7) | 4 (3.3) | 2 (1.7) | 4 (3.3) | 2 (1.7) |
| Back pain | 1 (0.8) | 1 (0.8) | 2 (1.7) | 2 (1.7) | 4 (3.3) | 2 (1.7) | 1 (0.8) | 1 (0.8) |
| Musculoskeletal chest pain | 1 (0.8) | 0 (0) | 0 (0) | 0 (0) | 0 (0) | 0 (0) | 0 (0) | 0 (0) |
| Musculoskeletal pain | 1 (0.8) | 1 (0.8) | 2 (1.7) | 1 (0.8) | 0 (0) | 0 (0) | 1 (0.8) | 0 (0) |
| Myalgia | 15 (12.5) | 18 (15.0) | 14 (11.8) | 15 (12.5) | 14 (11.7) | 22 (18.3) | 14 (11.7) | 16 (13.3) |
| **Nervous system disorders** | 24 (20.0) | 29 (24.2) | 30 (25.2) | 36 (30.0) | 33 (27.5) | 25 (20.8) | 30 (25.0) | 29 (24.2) |
| Dizziness | 6 (5.0) | 4 (3.3) | 3 (2.5) | 4 (3.3) | 4 (3.3) | 5 (4.2) | 3 (2.5) | 2 (1.7) |
| Headache | 21 (17.5) | 26 (21.7) | 27 (22.7) | 30 (25.0) | 30 (25.0) | 22 (18.3) | 29 (24.2) | 28 (23.3) |
| Hypoesthesia | 0 (0) | 0 (0) | 0 (0) | 0 (0) | 0 (0) | 0 (0) | 1 (0.8) | 0 (0) |
| Neuropathy peripheral | 0 (0) | 0 (0) | 0 (0) | 1 (0.8) | 0 (0) | 0 (0) | 0 (0) | 0 (0) |
| Paresthesia | 0 (0) | 1 (0.8) | 0 (0) | 1 (0.8) | 0 (0) | 1 (0.8) | 0 (0) | 0 (0) |
| Somnolence | 0 (0) | 1 (0.8) | 0 (0) | 0 (0) | 0 (0) | 0 (0) | 0 (0) | 0 (0) |
| **Psychiatric disorders** | 0 (0) | 0 (0) | 0 (0) | 0 (0) | 0 (0) | 0 (0) | 2 (1.7) | 0 (0) |
| Insomnia | 0 (0) | 0 (0) | 0 (0) | 0 (0) | 0 (0) | 0 (0) | 1 (0.8) | 0 (0) |
| Libido decreased | 0 (0) | 0 (0) | 0 (0) | 0 (0) | 0 (0) | 0 (0) | 1 (0.8) | 0 (0) |
| **Renal and urinary disorders** | 0 (0) | 0 (0) | 0 (0) | 0 (0) | 0 (0) | 1 (0.8) | 0 (0) | 0 (0) |
| Dysuria | 0 (0) | 0 (0) | 0 (0) | 0 (0) | 0 (0) | 1 (0.8) | 0 (0) | 0 (0) |
| **Reproductive system and breast disorders** | 0 (0) | 2 (1.7) | 0 (0) | 2 (1.7) | 0 (0) | 1 (0.8) | 2 (1.7) | 1 (0.8) |
| Breast inflammation | 0 (0) | 0 (0) | 0 (0) | 0 (0) | 0 (0) | 0 (0) | 1 (0.8) | 0 (0) |
| Dysfunctional uterine bleeding | 0 (0) | 1 (0.8) | 0 (0) | 0 (0) | 0 (0) | 0 (0) | 0 (0) | 0 (0) |
| Dysmenorrhea | 0 (0) | 0 (0) | 0 (0) | 1 (0.8) | 0 (0) | 0 (0) | 0 (0) | 1 (0.8) |
| Erectile dysfunction | 0 (0) | 1 (0.8) | 0 (0) | 0 (0) | 0 (0) | 0 (0) | 0 (0) | 0 (0) |
| Genital ulceration | 0 (0) | 0 (0) | 0 (0) | 0 (0) | 0 (0) | 1 (0.8) | 1 (0.8) | 0 (0) |
| Uterine hemorrhage | 0 (0) | 0 (0) | 0 (0) | 1 (0.8) | 0 (0) | 0 (0) | 0 (0) | 0 (0) |
| **Respiratory, thoracic and mediastinal disorders** | 9 (7.5) | 5 (4.2) | 6 (5.0) | 7 (5.8) | 7 (5.8) | 9 (7.5) | 3 (2.5) | 10 (8.3) |
| Asthma | 0 (0) | 0 (0) | 1 (0.8) | 0 (0) | 0 (0) | 0 (0) | 0 (0) | 1 (0.8) |
| Cough | 4 (3.3) | 3 (2.5) | 2 (1.7) | 0 (0) | 3 (2.5) | 7 (5.8) | 1 (0.8) | 3 (2.5) |
| Epistaxis | 0 (0) | 0 (0) | 0 (0) | 1 (0.8) | 0 (0) | 0 (0) | 0 (0) | 0 (0) |
| Productive cough | 0 (0) | 0 (0) | 1 (0.8) | 0 (0) | 0 (0) | 0 (0) | 0 (0) | 0 (0) |
| Rhinitis allergic | 0 (0) | 1 (0.8) | 0 (0) | 2 (1.7) | 1 (0.8) | 0 (0) | 0 (0) | 2 (1.7) |
| Rhinorrhea | 5 (4.2) | 1 (0.8) | 3 (2.5) | 3 (2.5) | 3 (2.5) | 1 (0.8) | 2 (1.7) | 4 (3.3) |
| Throat irritation | 0 (0) | 0 (0) | 0 (0) | 0 (0) | 0 (0) | 1 (0.8) | 0 (0) | 0 (0) |
| Tonsillar inflammation | 0 (0) | 0 (0) | 0 (0) | 1 (0.8) | 0 (0) | 0 (0) | 0 (0) | 0 (0) |
| **Skin and subcutaneous tissue disorders** | 3 (2.5) | 2 (1.7) | 5 (4.2) | 4 (3.3) | 3 (2.5) | 3 (2.5) | 3 (2.5) | 5 (4.2) |
| Dermatitis | 1 (0.8) | 0 (0) | 1 (0.8) | 2 (1.7) | 0 (0) | 0 (0) | 0 (0) | 2 (1.7) |
| Dermatitis allergic | 2 (1.7) | 0 (0) | 2 (1.7) | 0 (0) | 1 (0.8) | 2 (1.7) | 1 (0.8) | 0 (0) |
| Dermatitis contact | 0 (0) | 0 (0) | 0 (0) | 0 (0) | 0 (0) | 0 (0) | 1 (0.8) | 0 (0) |
| Itching scar | 0 (0) | 1 (0.8) | 0 (0) | 0 (0) | 0 (0) | 0 (0) | 0 (0) | 0 (0) |
| Night sweats | 0 (0) | 0 (0) | 0 (0) | 0 (0) | 0 (0) | 0 (0) | 1 (0.8) | 0 (0) |
| Pruritus | 0 (0) | 1 (0.8) | 2 (1.7) | 0 (0) | 0 (0) | 1 (0.8) | 0 (0) | 3 (2.5) |
| Pruritus generalized | 0 (0) | 0 (0) | 0 (0) | 1 (0.8) | 0 (0) | 0 (0) | 0 (0) | 0 (0) |
| Skin irritation | 0 (0) | 0 (0) | 0 (0) | 0 (0) | 1 (0.8) | 0 (0) | 0 (0) | 0 (0) |
| Skin lesion | 0 (0) | 0 (0) | 0 (0) | 0 (0) | 1 (0.8) | 0 (0) | 0 (0) | 0 (0) |

| | | | | | | | | |
|---|---|---|---|---|---|---|---|---|
| Urticaria | 0 (0) | 0 (0) | 0 (0) | 1 (0.8) | 0 (0) | 0 (0) | 0 (0) | 0 (0) |

*MedDRA version 20.0*

Table S14: Severity of adverse events up to Day 28 post-vaccination by study arm, and vaccine manufacturer, safety population

| **Severity** | **Bio-Manguinhos** | | **Chumakov IPVE** | | **IPD** | | **Sanofi Pasteur** | |
|---|---|---|---|---|---|---|---|---|
| n (%) with ≥ 1 AE | Frac. N=120 | Standard N=120 | Frac. N=119 | Standard N=120 | Frac. N=120 | Standard N=120 | Frac. N=120 | Standard N=120 |
| Mild | 57 (47.5) | 66 (55.0) | 65 (54.6) | 67 (55.8) | 73 (60.8) | 69 (57.5) | 54 (45.0) | 69 (57.5) |
| Moderate | 4 (3.3) | 7 (5.8) | 8 (6.7) | 11 (9.2) | 9 (7.5) | 6 (5.0) | 10 (8.3) | 9 (7.5) |
| Severe | 0 (0) | 0 (0) | 0 (0) | 0 (0) | 0 (0) | 0 (0) | 0 (0) | 0 (0) |
| Life Threatening | 0 (0) | 0 (0) | 0 (0) | 0 (0) | 0 (0) | 0 (0) | 0 (0) | 0 (0) |

Table S15: Proportion of participants with at least one related event up to Day 28 post-vaccination, by study arm, and vaccine manufacturer, safety population

| **Relatedness** | **Bio-Manguinhos** | | **Chumakov IPVE** | | **IPD** | | **Sanofi Pasteur** | |
|---|---|---|---|---|---|---|---|---|
| n (%) with ≥ 1 AE | Frac. N=120 | Standard N=120 | Frac. N=119 | Standard N=120 | Frac. N=120 | Standard N=120 | Frac. N=120 | Standard N=120 |
| Related | 38 (31.7) | 50 (41.7) | 55 (46.2) | 55 (45.8) | 59 (49.2) | 50 (41.7) | 48 (40.0) | 53 (44.2) |

Table S16: Outcome of adverse events up to Day 28 post-vaccination, by study arm, and vaccine manufacturer, safety population

| **Outcome** | **Bio-Manguinhos** | | **Chumakov IPVE** | | **IPD** | | **Sanofi Pasteur** | |
|---|---|---|---|---|---|---|---|---|
| n (%) with ≥ 1 AE | Frac. N=120 | Standard N=120 | Frac. N=119 | Standard N=120 | Frac. N=120 | Standard N=120 | Frac. N=120 | Standard N=120 |
| Recovered | 58 (48.3) | 69 (57.5) | 69 (58.0) | 72 (60.0) | 76 (63.3) | 72 (60.0) | 60 (50.0) | 70 (58.3) |
| Recovered with sequelae | 0 (0) | 0 (0) | 0 (0) | 0 (0) | 0 (0) | 0 (0) | 0 (0) | 0 (0) |

### Serious Adverse Events

Table S17: Summary of serious adverse events by study arm, and vaccine manufacturer, safety population

| | Bio-Manguinhos | | Chumakov IPVE | | IPD | | Sanofi Pasteur | |
|---|---|---|---|---|---|---|---|---|
| | Frac. N=120 | Standard N=120 | Frac. N=119 | Standard N=120 | Frac. N=120 | Standard N=120 | Frac. N=120 | Standard N=120 |
| n (%) with ≥ 1 SAE | 2 (1.7) | 1 (0.8) | 0 (0) | 0 (0) | 3 (2.5) | 2 (1.7) | 0 (0) | 2 (1.7) |

Table S18: Serious adverse events by MedDRA (v20.0) coding by study arm and vaccine manufacturer, safety population

| System Organ Class and Preferred Term | Bio-Manguinhos | | Chumakov IPVE | | IPD | | Sanofi Pasteur | |
|---|---|---|---|---|---|---|---|---|
| n (%) with ≥ 1 SAE | Frac. N=120 | Standard N=120 | Frac. N=119 | Standard N=120 | Frac. N=120 | Standard N=120 | Frac. N=120 | Standard N=120 |
| **General disorders and administration site conditions** | 0 (0) | 0 (0) | 0 (0) | 0 (0) | 0 (0) | 1 (0.8) | 0 (0) | 0 (0) |
| Sudden death | 0 (0) | 0 (0) | 0 (0) | 0 (0) | 0 (0) | 1 (0.8) | 0 (0) | 0 (0) |
| **Infections and infestations** | 1 (0.8) | 0 (0) | 0 (0) | 0 (0) | 0 (0) | 1 (0.8) | 0 (0) | 1 (0.8) |
| Malaria | 1 (0.8) | 0 (0) | 0 (0) | 0 (0) | 0 (0) | 1 (0.8) | 0 (0) | 0 (0) |
| Wound sepsis | 0 (0) | 0 (0) | 0 (0) | 0 (0) | 0 (0) | 0 (0) | 0 (0) | 1 (0.8) |
| **Injury, poisoning, and procedural complaints** | 0 (0) | 0 (0) | 0 (0) | 0 (0) | 1 (0.8) | 0 (0) | 0 (0) | 0 (0) |
| Road traffic accident | 0 (0) | 0 (0) | 0 (0) | 0 (0) | 1 (0.8) | 0 (0) | 0 (0) | 0 (0) |
| **Pregnancy, puerperium and perinatal conditions** | 0 (0) | 0 (0) | 0 (0) | 0 (0) | 1 (0.8) | 0 (0) | 0 (0) | 0 (0) |
| Spontaneous incomplete abortion | 0 (0) | 0 (0) | 0 (0) | 0 (0) | 1 (0.8) | 0 (0) | 0 (0) | 0 (0) |
| **Respiratory, thoracic and mediastinal disorders** | 0 (0) | 1 (0.8) | 0 (0) | 0 (0) | 0 (0) | 0 (0) | 0 (0) | 0 (0) |
| Lung neoplasm malignant | 0 (0) | 1 (0.8) | 0 (0) | 0 (0) | 0 (0) | 0 (0) | 0 (0) | 0 (0) |
| **Surgical and medical procedures** | 1 (0.8) | 0 (0) | 0 (0) | 0 (0) | 1 (0.8) | 0 (0) | 0 (0) | 1 (0.8) |
| Caesarean section | 1 (0.8) | 0 (0) | 0 (0) | 0 (0) | 1 (0.8) | 0 (0) | 0 (0) | 1 (0.8) |

*MedDRA version 20.0*

Table S19: Outcome of serious adverse events, by study arm, and vaccine manufacturer, safety population

| Outcome | Bio-Manguinhos | | Chumakov IPVE | | IPD | | Sanofi Pasteur | |
|---|---|---|---|---|---|---|---|---|
| n (%) with ≥ 1 SAE | Frac. N=120 | Standard N=120 | Frac. N=119 | Standard N=120 | Frac. N=120 | Standard N=120 | Frac. N=120 | Standard N=120 |
| Recovered | 2 (1.7) | 0 (0) | 0 (0) | 0 (0) | 2 (1.7) | 1 (0.8) | 0 (0) | 1 (0.8) |
| Recovered with sequelae | 0 (0) | 0 (0) | 0 (0) | 0 (0) | 0 (0) | 0 (0) | 0 (0) | 1 (0.8) |
| Fatal or died | 0 (0) | 1 (0.8) | 0 (0) | 0 (0) | 1 (0.8) | 1 (0.8) | 0 (0) | 0 (0) |
